# The Socio-economic Shield Limits Lassa Virus Spillover in Urban West Africa

**DOI:** 10.64898/2025.12.17.25342147

**Authors:** David Simons

**Affiliations:** Department of Anthropology, Pennsylvania State University, State College, USA

**Keywords:** Lassa virus, Spillover, Joint Species Distribution Model, Mastomys natalensis, Seroreversion, Underreporting, Urbanisation

## Abstract

Spatial risk models for Lassa fever (LF) generally predict the primary reservoir, *Mastomys natalensis*, is restricted to rural landscapes. This study integrates multi-species biotic interactions and anthropogenic land-use into a high-resolution framework to evaluate LF’s urban potential. I implemented an Integrated Multi-Species Occupancy Model (IMSOM) to reconstruct the reservoir’s realised niche, accounting for sampling bias and invasive rodent competitors. A socio-economic filter, proxied by nighttime lights, was introduced to model the dampening effect of urban infrastructure on spillover. Annual infections were estimated using a demographic compartmental model incorporating empirical seroreversion rates. Results indicate high biological hazard across the peri-urban fringes of major West African cities. However, an infrastructure-driven socio-economic shield decouples this hazard from human incidence in dense urban cores. Accounting for spatial shielding and antibody waning yields an estimated 2.6 million annual LASV infections. Comparing predictions to clinical data reveals substantial surveillance gaps, identifying highly suitable silent districts in Nigeria, Benin, and Togo with zero reported cases. LF possesses the biological potential to become a peri-urban disease; addressing these surveillance gaps at the peri-urban interface is a critical public health priority.

**Key Findings:**

- Accounting for species interactions reveals that the Lassa fever reservoir (M. natalensis) exhibits high ecological tolerance for peri-urban and human-modified landscapes.
- Urban infrastructure acts as a socio-economic shield, decoupling biological hazard from realised human spillover in densely populated city centres.
- Adjusting for realistic antibody waning and urban shielding yields an estimated regional burden of 2.6 million annual Lassa virus infections.
- Spatial validation identifies numerous silent districts across West Africa, revealing surveillance gaps rather than an absence of biological hazard.

## 1. Introduction

The rapid urbanisation of the tropics presents a critical ecological paradox for zoonotic disease risk. While high human densities and the proliferation of commensal reservoirs theoretically increase contact rates, urban infrastructure may act as a selective transmission filter. Resolving this tension is critical for forecasting the future of viral haemorrhagic fevers in the Anthropocene. Lassa fever (LF), caused by *Mammarenavirus lassaense* (LASV; Arenaviridae), represents an ideal system to test these dynamics. Its reservoir, the generalist commensal *Mastomys natalensis*, is having its realised niche actively reshaped by the interaction between urban expansion and competitive pressure from invasive rodents.

LF is a significant public health concern across West Africa, yet accurate estimation of the true disease burden remains elusive due to high rates of asymptomatic infection and persistent gaps in diagnostic surveillance [1–3]. Robust, spatially explicit risk maps are therefore essential for guiding public health interventions, including the deployment of future vaccines [4,5]. Previous forecasting efforts have successfully utilised mechanistic models to predict environmental suitability for the host (**D_M_**) and the virus (**D_L_**), calibrating the composite hazard (**D**_**X**_ = **D**_**M**_ × **D**_**L**_) against human seroprevalence data [1]. However, these foundational models relied primarily on abiotic predictors, predicting the climatic exclusion of the reservoir from highly modified urban landscapes.

In West Africa’s heterogeneous landscapes, the reservoir’s realised niche is constrained by biotic interactions [6]. In high-density urban environments, *M. natalensis* faces increasing competitive pressure from invasive commensals, specifically the Black Rat (*Rattus rattus*) and House Mouse (*Mus musculus*) [7]. By neglecting these biotic constraints, previous models have potentially mischaracterised the zoonotic hazard in cities, creating an urban blind spot in risk forecasting.

Furthermore, spillover is not solely a function of reservoir density, but of interface permeability [8]. In rural landscapes, porous housing materials create a high-contact interface [9]. Previous epidemiological frameworks, trained heavily on rural data, have projected these high-contact dynamics into dense cities. In reality, urbanisation introduces structural barriers including; improved housing, distinct agricultural zoning, and organised refuse management that act to decouple human exposure from rodent density [10]. Models that fail to account for this non-linear ‘Socio-economic Shield’ systematically overestimate spillover in urban centres, potentially directing surveillance resources away from the peri-urban fringes where transmission intensity is highest.

To address this gap, I re-evaluated the spatial hazard of LASV spillover using an Integrated Multi-Species Occupancy Model (IMSOM) [11,12]. This framework explicitly quantifies co-occurrence dynamics between *M. natalensis* and the wider rodent community, deriving a refined reservoir layer (**D_M_**) that accounts for the biotic pressure exerted by competitors.

To resolve the substantial uncertainty surrounding the magnitude of human infection, I integrated recent longitudinal evidence on LASV seroreversion rates to constrain the annual force of infection [13]. Finally, these refined estimates were validated against sub-national clinical case reports to quantify the discrepancy between biological hazard and passive surveillance, spatially identifying silent districts to guide the prioritisation of future LF control.

## 2. Methods

### 2.1. Data Assembly and Environmental Predictors

#### 2.1.1. Host Community and Spatial Framework

This study focused on the endemic zone of the A-I clade of *M. natalensis* in West Africa, evaluated across a 0.05^°^ × 0.05^°^ spatial grid. To characterise the host community, a comprehensive dataset of small mammal occurrences (1972–2022) was compiled from curated trapping databases, targeted literature extraction, and opportunistic records [14–16]. The dataset included eight epidemiologically and ecologically relevant species: the primary reservoir (*M. natalensis*), sympatric native species, and invasive commensals (*R. rattus, M. musculus*).

#### 2.1.2 Environmental Covariates

I reconstructed the environmental predictor stack utilised in previous forecasting efforts, updating the temporal window to 2001–2025 [1]. Predictors captured climate, topography, seasonal environmental stability, and anthropogenic land-use (details in Supplementary Methods). To prevent multicollinearity and ensure ecological parsimony, a final subset of predictors was selected based on Variance Inflation Factors (VIF < 3) and *a priori* biological hypotheses. This subset included indices of temperature, precipitation, elevation, agricultural density, and a quadratic term for log-transformed human population density to capture non-linear responses to urbanisation.

### 2.2. Ecological Modelling: The Realised Host Niche (D_M_)

To estimate the spatial hazard of LASV, I first refined the predicted distribution of the reservoir host (**D_M_**). Previous models relied on abiotic climatic envelopes. However, to account for the competitive exclusion of *M. natalensis* by invasive rodents in urban environments, I implemented an Integrated Multi-Species Occupancy Model (IMSOM) using the `spOccupancy` package in R [11,17].

Unlike single-species approaches, the IMSOM explicitly models residual correlations between species via latent factor analysis, leveraging community-level data pooling to stabilise detection estimates for unstructured opportunistic records. This joint framework is essential for the subsequent pathogen modelling; it concurrently generates the predicted spatial distributions of invasive competitors (*R. rattus, M. musculus*), which is a prerequisite for testing the biotic constraints on viral prevalence. While random background pseudo-absences were generated to characterise available environmental space, spatial sampling bias was controlled for within the model’s observation process, which parameterised detection probability as a function of data source and survey effort (Supplementary Methods).

From the fitted model, I extracted the mean posterior occupancy probability for *M. natalensis* to serve as the refined reservoir hazard layer (**D_M_**), masked to its IUCN extant range to prevent biogeographical extrapolation. I concurrently extracted the predicted occupancy surfaces for the invasive competitors *R. rattus* and *M. musculus*. Given their low observed rates of active LASV infection and unproven competence for onward transmission, these invasive species were treated primarily as ecological constraints on the reservoir, rather than direct sources of zoonotic hazard. Their occupancy layers were subsequently integrated as biotic covariates in the pathogen model.

### 2.3. Spillover Estimation and Epidemiological Constraints

#### 2.3.1. Modelling the Biotic and Socio-economic Filters

The composite ecological hazard (**D_X_**) was calculated as the product of the reservoir distribution (**D_M_**) and pathogen prevalence (**D_L_**). To construct **D_L_**, I trained a Boosted Regression Tree (BRT) on 73 testing sites where *M. natalensis* was assayed for LASV. To explicitly account for the hypothesised dilution effect, predicted occupancies of invasive rodents (*R. rattus, M. musculus*) from the IMSOM were included as biotic covariates. To address spatial circularity, any rodent testing site within 5 km of a human serosurvey used for final calibration was excluded from the training set.

Unlike previous models, I explicitly introduced a socio-economic filter using nighttime lights (NTL) to proxy electrification and housing quality, hypothesising that urban infrastructure acts as a physical barrier to rodent contact. Because empirical community-based serosurveys are entirely absent from high-density city centres [18], correlative models risk extrapolating high rural contact rates into novel urban environments. To prevent this ecological fallacy, I augmented the calibration dataset with five synthetic absence anchors (*n* = 500, 0% seroprevalence) representing hyper-urban commercial centres (e.g., Lagos Island). These act as informative Bayesian priors reflecting the historical absence of autochthonous transmission in these zones, constraining the model to estimate the suppressive effect of extreme urbanisation (NTL>20).

#### 2.3.2. Transmission Dynamics and Incidence Estimation

The composite ecological hazard (**D_X_**) was calibrated against 94 historical human serosurveys using a quasi-binomial Generalised Linear Model (GLM). To convert predicted equilibrium seroprevalence (*Ω* ^*^) into the annual incidence of new infections, I utilised a steady-state demographic Susceptible-Infectious-Recovered-Susceptible (SIRS) compartmental model [1].

At endemic steady-state, the incidence of new infections must balance the total outflow of individuals recovering and dying (Full system of Ordinary Differential Equations provided in Supplementary Methods). By solving this system at equilibrium, the total annual incidence of new infections within a given spatial cell can be derived directly from the observed seropositive fraction (*Ω*^*^) and the total population (*N* ):

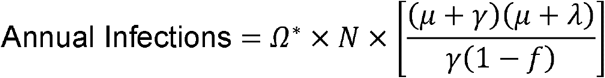

This formulation addresses the demographic reality that a fraction of infected individuals die before seroconverting, and that surviving seropositive individuals are continuously removed via natural demographic turnover (*μ*).

To capture regional demographic heterogeneity, the background mortality rate (*μ*) was treated as a spatially varying covariate, calculated as the reciprocal of country-specific life expectancies derived from 2023 estimates (ranging from 54 years in Nigeria to 69 years in Senegal). The recovery rate (*γ*) was set to 12 year^−1^, approximating a one-month infectious and convalescent period [13]. The infection fatality rate (*f*) was conservatively set to 2% (0.02) [2].

Uncertainty surrounding the total burden was constrained by evaluating a sensitivity range for the rate of antibody waning (*λ*). Relying on longitudinal serological evidence, I calculated the primary incidence burden assuming an annual seroreversion rate of 3%(*λ* = 0.03), with sensitivity bounds between lifelong immunity (*λ* = 0) and rapid loss (*λ* = 0.064). Total annual infections per 0.05^°^ pixel were calculated by integrating these parameters with the WorldPop 2020 human population matrix.

#### 2.3.3. Validation and Urban Gradient Analysis

To assess epidemiological accuracy, I compared predicted annual infections against aggregated clinical case reports at sub-national (Admin-2) and national scales [19]. I distinguish between total biological transmission events (predicted infections) and the subset of symptomatic, health-seeking individuals (reported cases). Rather than treating discrepancies purely as model error, spatial divergence was utilised to identify silent districts, areas where environmental suitability implies high transmission despite zero reported cases.

Second, to validate the ecological realism of the model within highly modified environments, I formally tested the ‘Urban Paradox’, the hypothesis that infrastructure creates a transmission blind spot in urban cores despite high biological hazard. I conducted a spatially explicit gradient analysis along 50 km radial transects for 104 stratified West African settlements. By extracting mean values for Ecological Hazard (**D_X_**), the Socio-economic Shield (NTL), and Realised Incidence across these transects, I quantified the spatial decoupling of biological threat and human exposure across different urban typologies (Full extraction protocols and statistical methods detailed in Supplementary Methods).

## 3. Results

### 3.1. Model Performance and the Cryptic Reservoir Niche

Current understanding of the LASV reservoir niche is constrained by severe sampling bias. Analysis of 4,908 unique rodent trapping locations revealed that 67% of systematic sampling effort (and 95% of opportunistic recording) has been concentrated in rural settings, leaving substantial spatial sampling gaps in the rapidly urbanising coastal corridors of West Africa (Supplementary Table 1; Figure 1). However, of the 228 systematic surveys successfully conducted in urban settings, 88.6% reported *M. natalensis* presence, suggesting the reservoir routinely exploits these environments.

**Figure 1.**
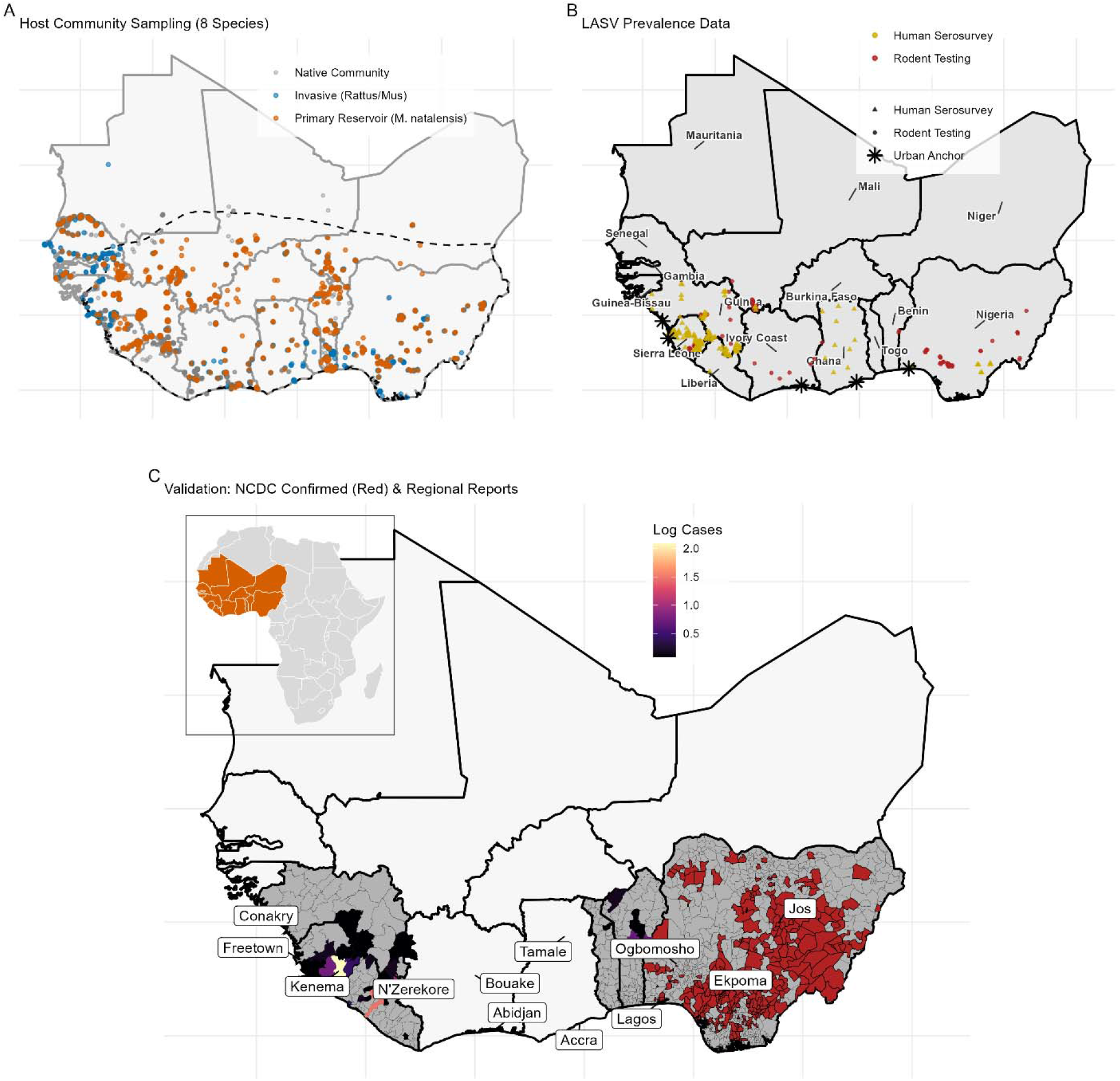
Spatial distribution of biological and epidemiological data. (A) Small mammal trapping locations used to train the IMSOM, stratified by ecological guild: the primary reservoir Mastomys natalensis (orange), invasive competitors *R. rattus*/*M. musculus* (blue), and the wider native community (grey). The dashed black line represents the IUCN range of *M. natalensis* within West Africa. (B) LASV prevalence data points. Red circles indicate rodent testing sites (PCR/Serology); gold triangles indicate human serosurveys used for calibration; black asterisks indicate synthetic urban absence anchors added to constrain model predictions in high-density city centres. (C) Reported cases resolved to administrative level 2 areas. Red LGAs in Nigeria indicate those that have reported at least one confirmed case between 2018 and 2025 (obtained from weekly situation reports produced by the NCDC). Outside of Nigeria administrative level 2 areas are coloured by the number of cases (log transformed) reported in the period 2012-2022. Labels refer to the focal cities for the spatially explicit gradient analyses. The inset map shows the extent of the study area within Africa. **Alt-Text for Figure 1:** A three-panel map of West Africa detailing the spatial distribution of study data. Panel A shows rodent trapping locations, visually highlighting a dense clustering of sampling effort in rural inland areas compared to the urbanised coast. Panel B maps pathogen testing sites, showing human serosurveys and rodent testing locations scattered across the region, alongside newly added synthetic absence anchors in high-density coastal city centres. Panel C displays reported sub-national clinical cases, showing intense clustering of confirmed cases within Nigeria and the Mano River Union, with an inset map confirming the study’s location within the African continent.

To formally test whether anthropogenic factors drive reservoir distribution beyond standard climatic envelopes, I compared a baseline climate-only occupancy model (analogous to previous forecasting efforts) against a model incorporating human population density and land-use. The inclusion of anthropogenic predictors substantially improved model fit (*Δ*WAIC = 70.83), confirming that the reservoir’s spatial limits cannot be defined by climate alone.

To reconstruct this realised niche while accounting for species interactions, I utilised the Integrated Multi-Species Occupancy Model (IMSOM). Previous foundational models, relying strictly on abiotic predictors, explicitly predicted the reservoir to be less prevalent along the heavily modified coastal corridors of southern Nigeria and West Africa (Basinski et al. 2021b). The biotically-informed IMSOM corrects this urban blind spot, predicting high occupancy probabilities (*P* > 0.7) across peri-urban and urban zones in these exact coastal regions (Figure 2).

**Figure 2.**
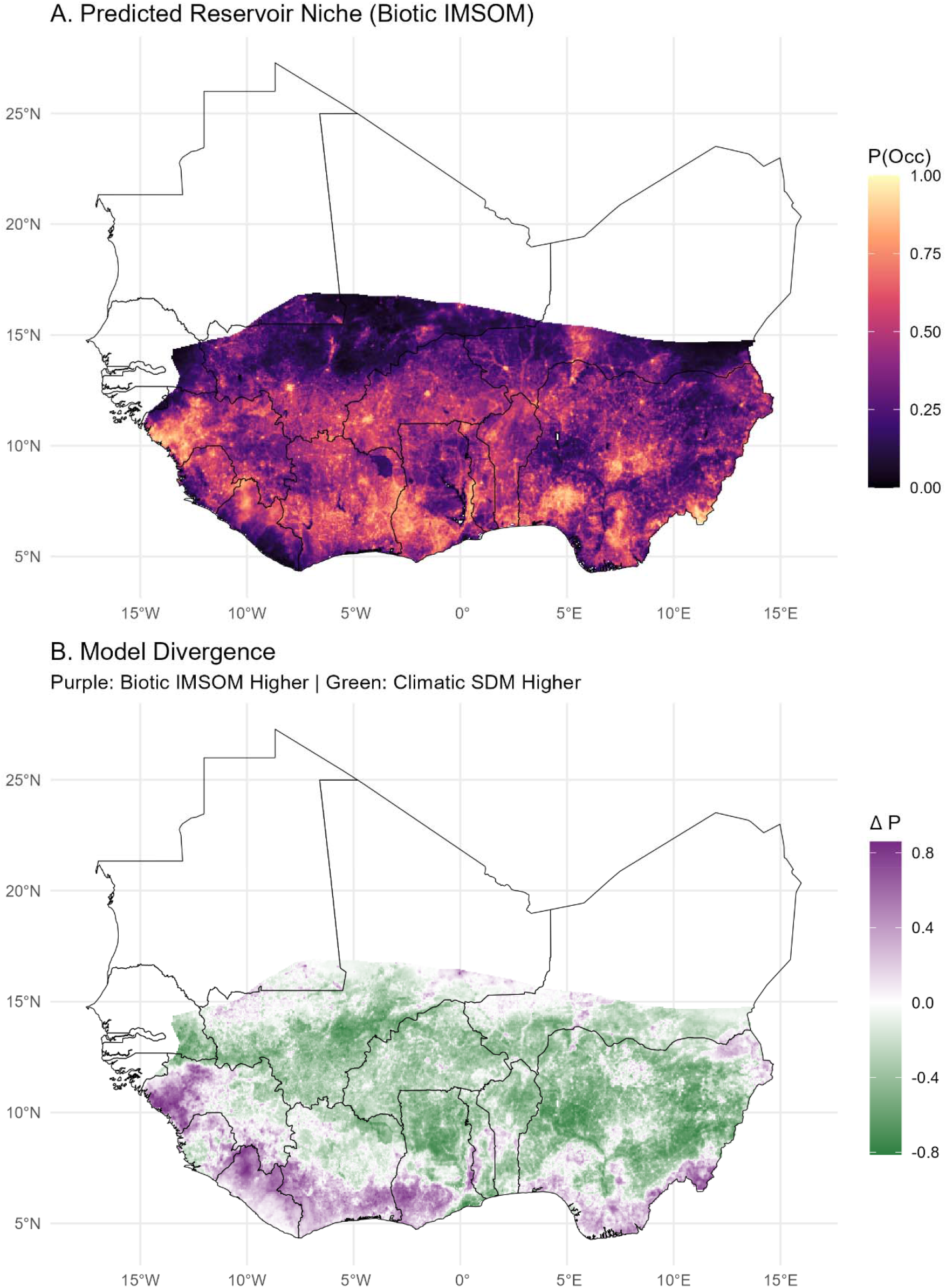
Predicted Reservoir Niche (*D*_*M*_). (A) IMSOM predicted occupancy probability for *M. natalensis*. (B) Difference map highlighting spatial divergence from previous climatic models, with positive values indicating higher predicted suitability in the IMSOM, particularly in urban zones. **Alt-Text for Figure 2:** A two-panel spatial map of West Africa. Panel A shows the predicted occupancy of Mastomys natalensis, revealing a strong latitudinal gradient where high ecological suitability is concentrated across the southern coastal countries (from Guinea to Nigeria), tapering off significantly in the northern Sahelian regions. Panel B maps the spatial difference between the new biotic model and older climatic models, visually demonstrating that the biotic model predicts substantially higher suitability along the heavily populated southern coastal corridor, whereas previous climatic models predicted higher suitability in drier, northern regions such as Senegal, Mali, and Burkina Faso.

This divergence is driven by the host’s empirically derived functional response to anthropogenic pressure. The IMSOM reveals a strong, positive association between *M. natalensis* occurrence and human population density (posterior mean *β* = 1.14, 95% Credible Interval [CrI]: 0.75–1.57). This synanthropic tolerance directly contrasts with native forest-specialist rodents like *Malacomys edwardsi*, which exhibited negative associations with anthropogenic disturbance (functional response coefficients for all species are shown in Supplementary Figure 1).

Spatially, this ecological tolerance manifests as an annular occupancy pattern in large megacities like Lagos. Rather than being restricted to the rural hinterland, the reservoir persists at high densities within informal settlements and peri-urban agricultural fringes (Figure 3; occupancy patterns for all 12 focal cities are provided in Supplementary Figures 2 and 3).

**Figure 3.**
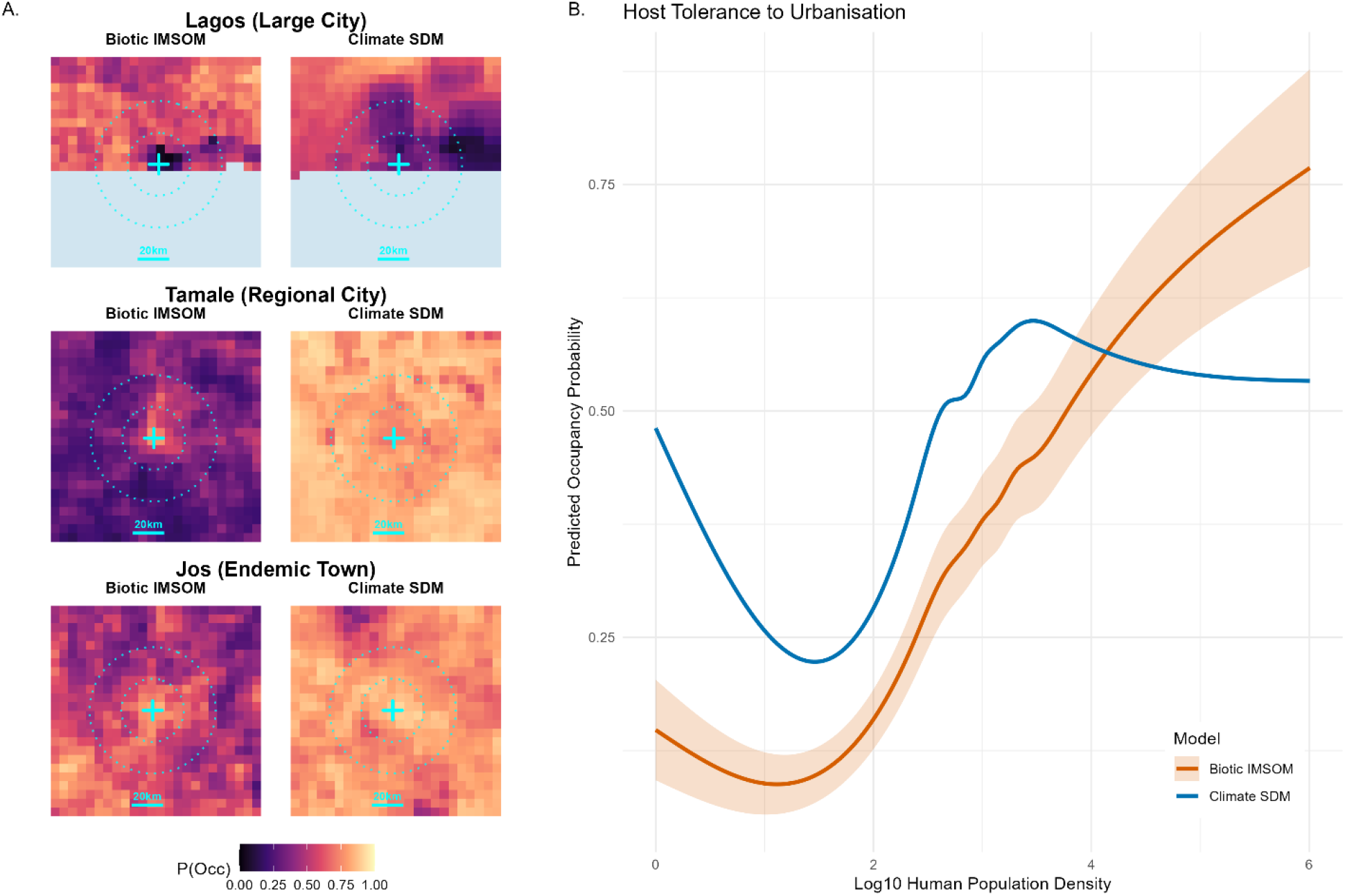
Mechanisms of Urban Tolerance. (A) Urban zooms showing reservoir persistence in peri-urban fringes for three example locations (Lagos, Nigeria; Tamale, Ghana and Jos, Nigeria. (B) Functional response curves of host occupancy to human population density for the climate based occupancy modelling (blue) and the IMSOM derived occupancy modelling (orange). **Alt-Text for Figure 3:** A two-part figure illustrating the reservoir’s tolerance to urbanisation. Panel A contains paired map zooms for three cities (Lagos, Tamale, and Jos), contrasting the older climatic model with the new biotic model. Visually, the biotic model predicts high reservoir occupancy extending deep into city centres and peri-urban fringes, correcting the ‘unsuitable’ voids predicted by the older climatic model. Panel B is a line graph comparing the functional response of host occupancy to increasing human population density. It highlights a stark divergence: while the climatic model’s predicted occupancy falls and plateaus at higher human densities, the biotic model’s curve climbs steeply and continuously upward into the highest population density extremes (up to a log10 density of 6), visually confirming the reservoir’s strong synanthropic tolerance.

### 3.2. Predictors of LASV Prevalence

Boosted Regression Tree (BRT) analysis indicates that Lassa virus prevalence within the reservoir is driven by a complex interplay of environmental stability and biotic interactions (Figure 4). While the strongest predictor was Greenness Persistence (NDVI Min, 19.0%), indicating a preference for habitats with stable vegetation cover—biotic interactions were highly influential. The predicted occupancy of the invasive black rat (*R. rattus*) was the second most important variable (17.7%), showing a strong positive association with viral prevalence in *M. natalensis*.

**Figure 4.**
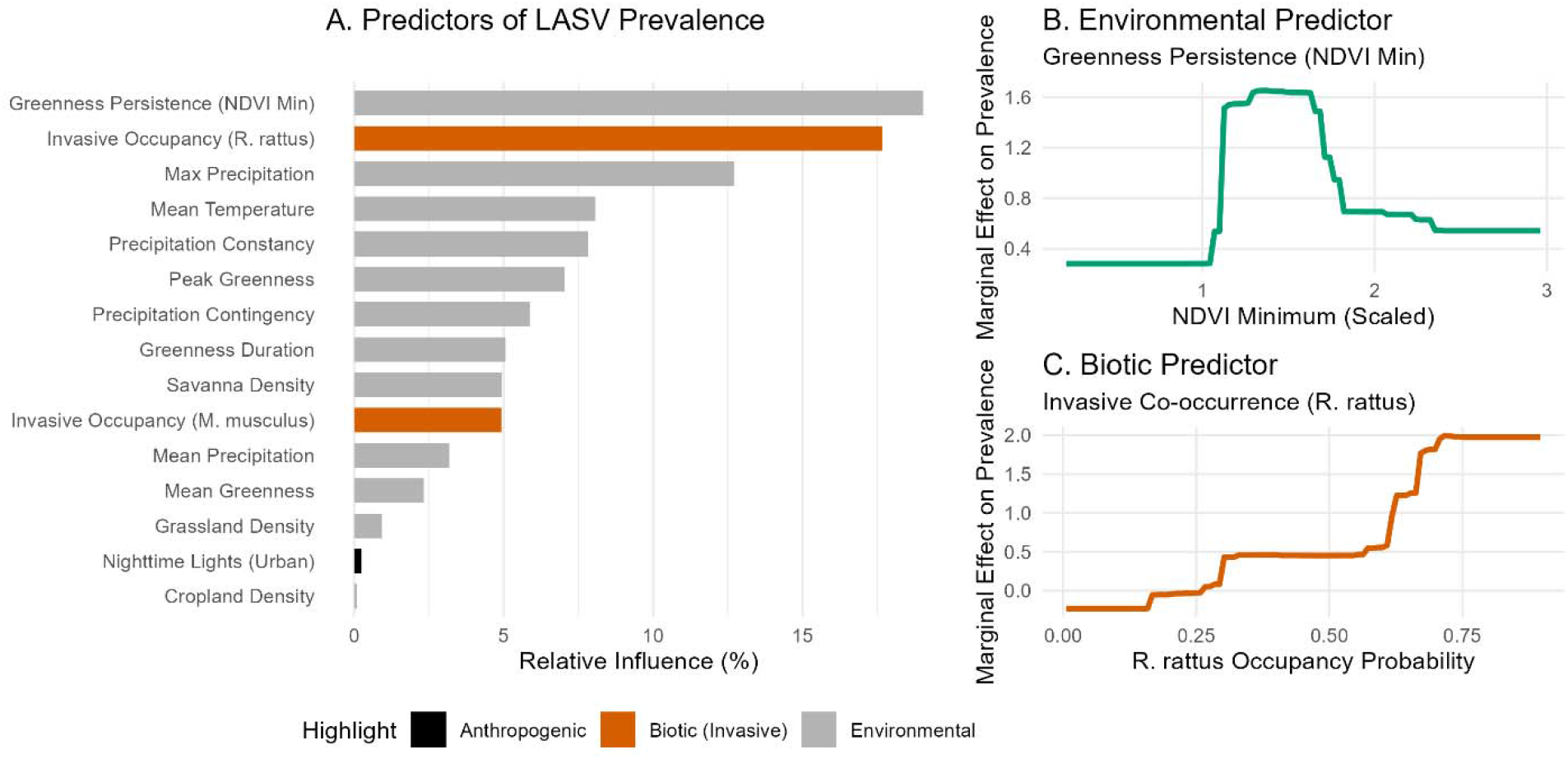
Predictors of Lassa Virus Prevalence. (A) Variable importance from the BRT model. (B) Partial dependence plot showing the non-linear association with greenness persistence (NDVI minimum). (C) Partial dependence plot showing the positive association with R. rattus occupancy. **Alt-Text for Figure 4:** A three-panel chart illustrating the ecological drivers of Lassa virus prevalence. Panel A is a bar chart ranking variable importance, showing that greenness persistence and invasive rodent occupancy are the two most influential predictors, outweighing standard temperature and precipitation metrics. Panel B is a line graph demonstrating a non-linear relationship where the marginal effect of greenness persistence on prevalence peaks strongly at intermediate vegetation levels before declining in the greenest environments. Panel C is a line graph displaying a strong positive association, where viral prevalence increases sharply in a step-like manner as the probability of Rattus rattus occupancy rises above 50 percent.

Bivariate partial dependence profiles (Supplementary Figure 4) reveal that peak viral prevalence does not occur in the most environmentally stable habitats (highest NDVI). Instead, it occupies a specific ecological niche characterised by intermediate greenness persistence, indicative of derived savannah or agricultural mosaics, co-occurring with high *R. rattus* occupancy.

The resulting biotically-informed Pathogen Hazard Layer (**D_L_**) indicates that viral circulation remains high in fragmented, human-dominated landscapes (Figure 5). Predicted viral suitability extends extensively across the derived savannah and peri-urban zones of Nigeria, Benin, and Togo, moving beyond the strict forest-margin zones emphasised in previous analyses.

**Figure 5.**
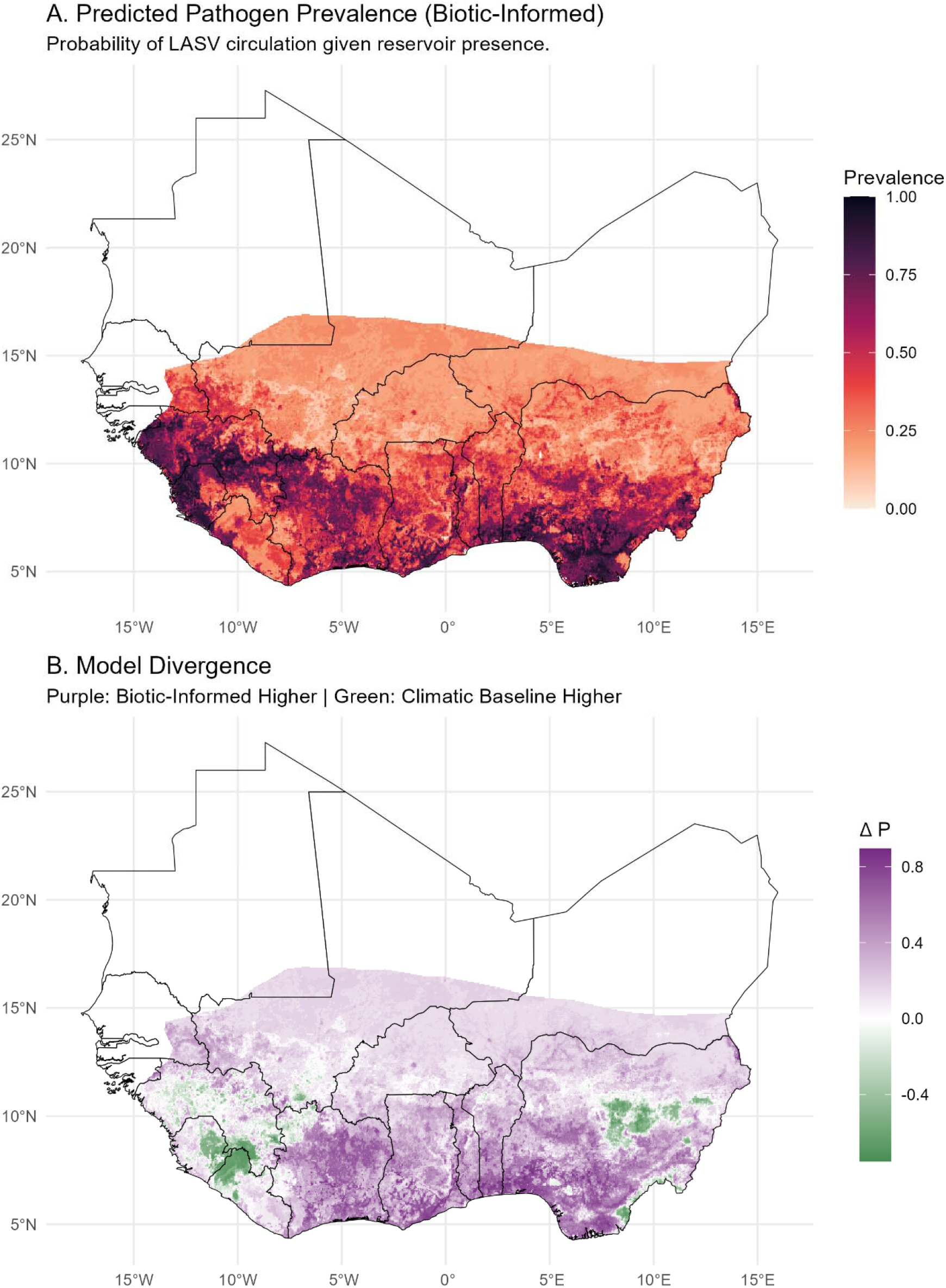
Predicted Pathogen Hazard (*D*_*L*_). Spatial distribution of viral prevalence in the reservoir host, conditional on reservoir presence. **Alt-Text for Figure 5:** A two-panel map of West Africa displaying the predicted spatial distribution of Lassa virus within the reservoir population. Panel A maps estimated pathogen prevalence, highlighting prominent hotspots of high viral circulation across Sierra Leone, Guinea, Southern Nigeria, the broader coastal regions, Southern Mali, and Northern Ivory Coast. Panel B is a difference map illustrating spatial divergences between the models. It visually demonstrates that the biotic-informed model predicts noticeably lower viral prevalence in specific focal areas, namely Eastern Sierra Leone, the forested border regions between Guinea and Liberia, and Nigeria’s Jos Plateau while remaining consistent with baseline predictions in other parts of Guinea and Sierra Leone.

This urban tolerance extends to the pathogen itself. While baseline climatic models predict a strict decline in viral prevalence as human population density increases (peaking at ∼2.5 log_10_ density before flattening), the biotically-informed model predicts a resurgence of viral risk at higher densities (log_10_ density ∼4–5). The predicted hazard is maintained in these dense transition zones before finally dropping in the most intensely urbanised commercial cores (Figure 6; urban zooms in Supplementary Figures 4 and 5).

**Figure 6.**
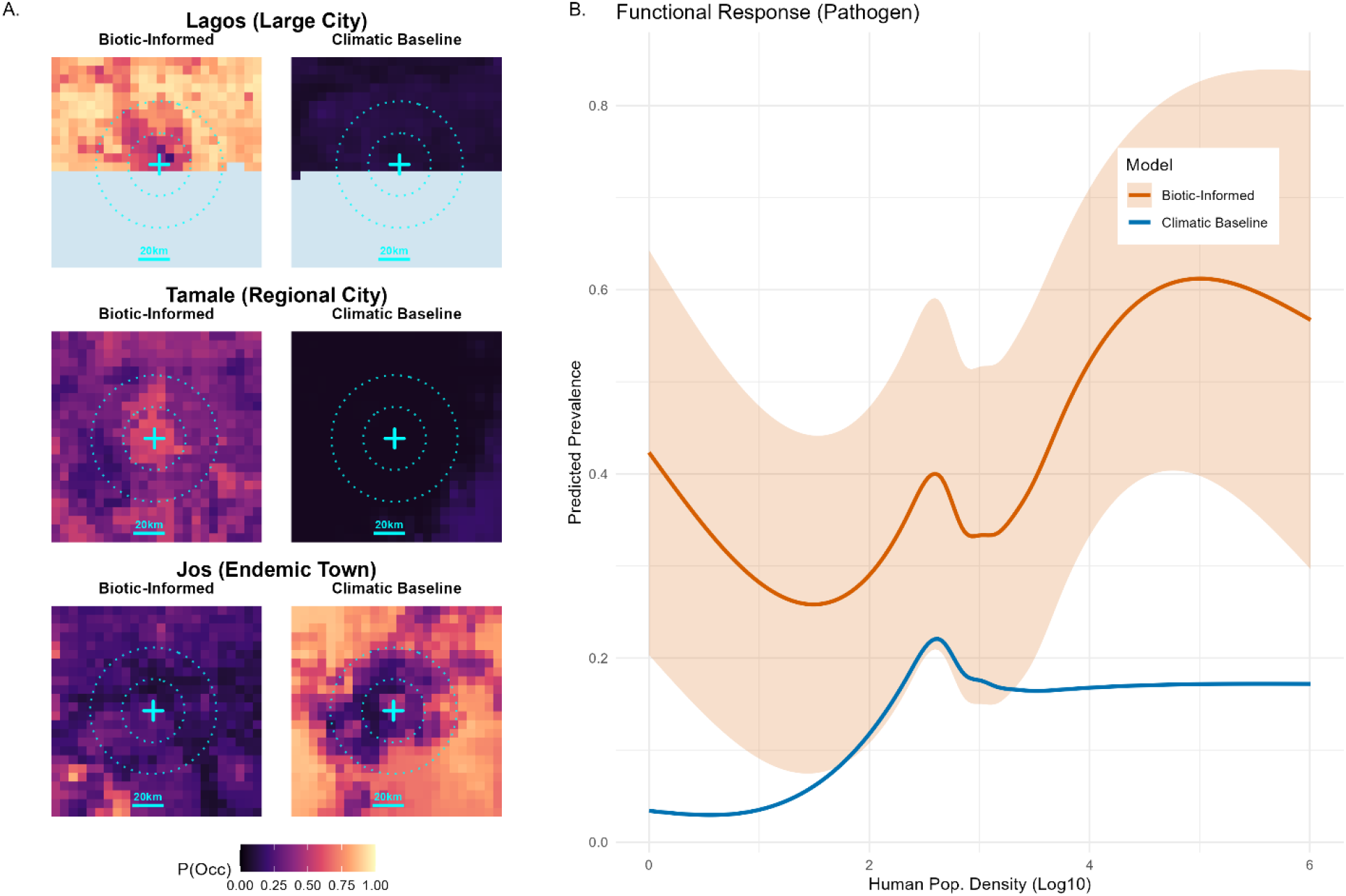
Pathogen Urban Tolerance. (A) Predicted viral prevalence (*D*_*L*_) in and around key cities, contrasting the biotic-informed model (left) with the climatic baseline (right). Contours indicate distance from city centre (20km). (B) Functional response of predicted prevalence to human population density (log_10_), showing the custom model’s (orange) persistence at higher densities compared to the climatic baseline (blue). **Alt-Text for Figure 6:** A two-part figure illustrating the tolerance of the Lassa virus to urbanisation. Panel A contains paired map zooms of predicted viral prevalence for three cities (Lagos, Tamale, and Jos). For Lagos and Tamale, the biotic model displays noticeably higher viral prevalence extending deeper into the urban centres compared to the older climatic baseline. For Jos, both models show a complex ring of peri-urban risk, though the biotic model predicts slightly lower overall probabilities. Panel B is a line graph comparing the functional response of viral prevalence to increasing human population density. While both models share an initial peak at lower densities, the climatic model sharply declines and flatlines as density increases. Conversely, the biotic model features a distinct second peak at high human population densities (around 5 on a log10 scale), visually demonstrating the pathogen’s persistence in highly populated transition zones.

### 3.3. The Socio-economic Shield and the Burden of Infection

To estimate spillover risk, I calculated the Composite Ecological Hazard (**D_X_** = **D_M_** × **D_X_**) However, despite this widespread ecological hazard, human infection patterns do not scale linearly with reservoir presence. I identified a non-linear Socio-economic Shield effect, where infrastructure quality (proxied by Nighttime Lights, NTL) dampens transmission efficiency.

Bootstrapped calibration curves demonstrate that at equivalent levels of Ecological Hazard (**D_X_**), human seroprevalence is significantly lower in urbanised settings compared to rural settings. This divergence is driven by a semi-mechanistic calibration: the urban curve is anchored by the limited available empirical urban serosurveys alongside synthetic absence priors representing hyper-urban cores, effectively forcing the model to acknowledge the historical absence of transmission in these highly developed zones. This decoupling is visually corroborated by the raw, un-binned data (Figure 7), which demonstrates how these heavily weighted urban anchors pull the expected seroprevalence strictly below the rural baseline.

**Figure 7.**
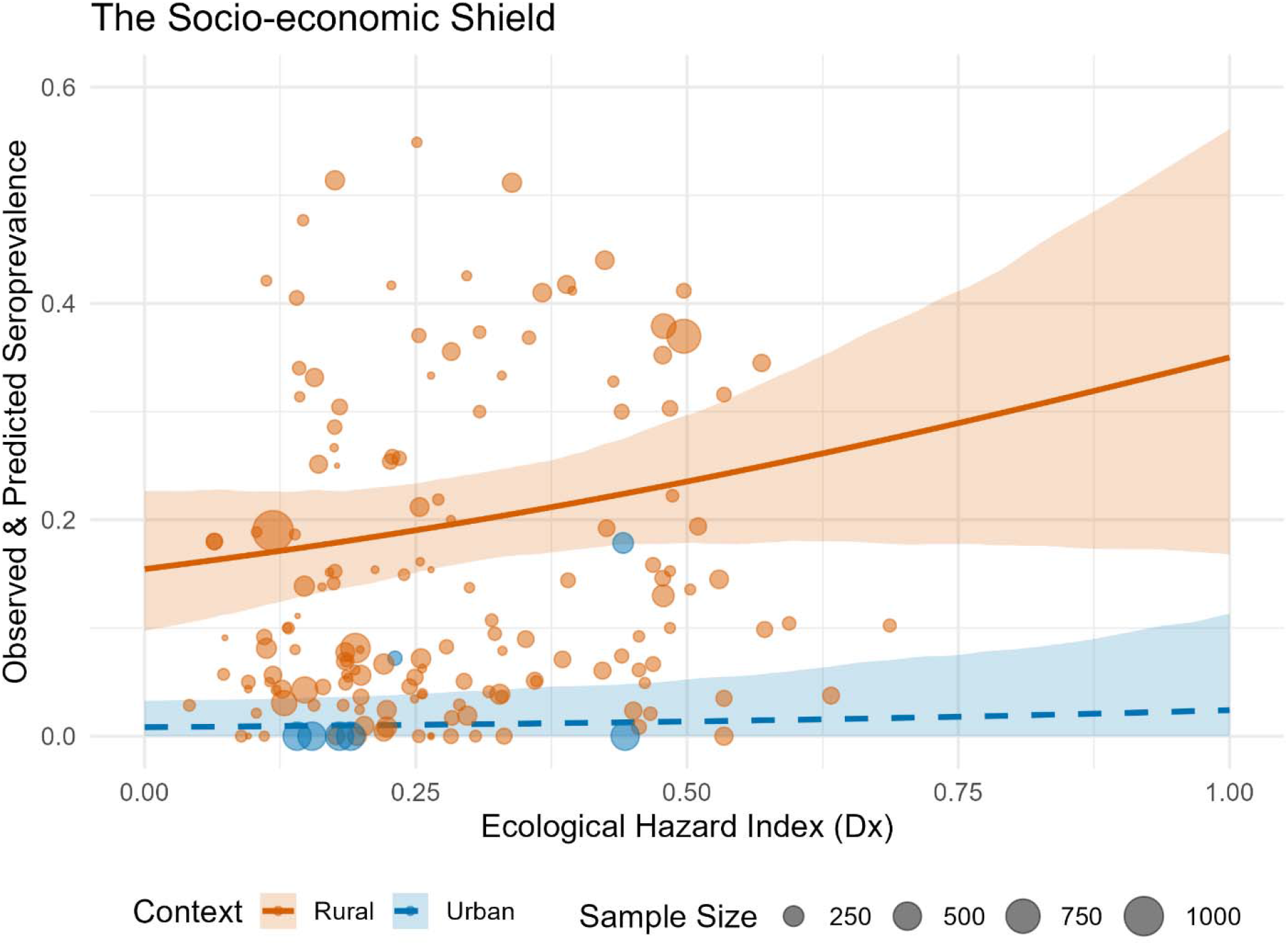
The Socio-economic Shield. Bootstrapped calibration curves showing the decoupling of ecological hazard and human seroprevalence in urban (blue) versus rural (orange) settings. Ribbons indicate 95% confidence intervals. Points indicate raw, un-binned observed seroprevalence from human serosurveys, with point size proportional to the total sample size of each study. **Alt-Text for Figure 7:** A line graph plotting observed human seroprevalence against predicted ecological hazard, stratified into rural and urban settings. The visual starkly demonstrates the socio-economic shield effect: the rural calibration curve shows a clear positive upward trend, rising from approximately 15 percent prevalence at zero hazard to 35 percent at maximum hazard. In sharp contrast, the urban calibration curve remains flat and near zero across all levels of ecological hazard. The plot also visually highlights a severe disparity in sampling data; the rural curve is supported by a wide scatter of highly variable empirical data points, whereas the urban curve relies on very few points that are overwhelmingly concentrated at zero prevalence.

The interplay between this Ecological Hazard and the Socio-economic Shield creates distinct transmission typologies characterised by strict spatial decoupling (Figure 8). To quantify this, radial risk profiles were extracted for a stratified sample of 104 West African settlements. Visualisation of the full profiles of ecological risk, infection incidence, and NTL are shown in Supplementary Figure 7 for the 12 focal cities.

**Figure 8.**
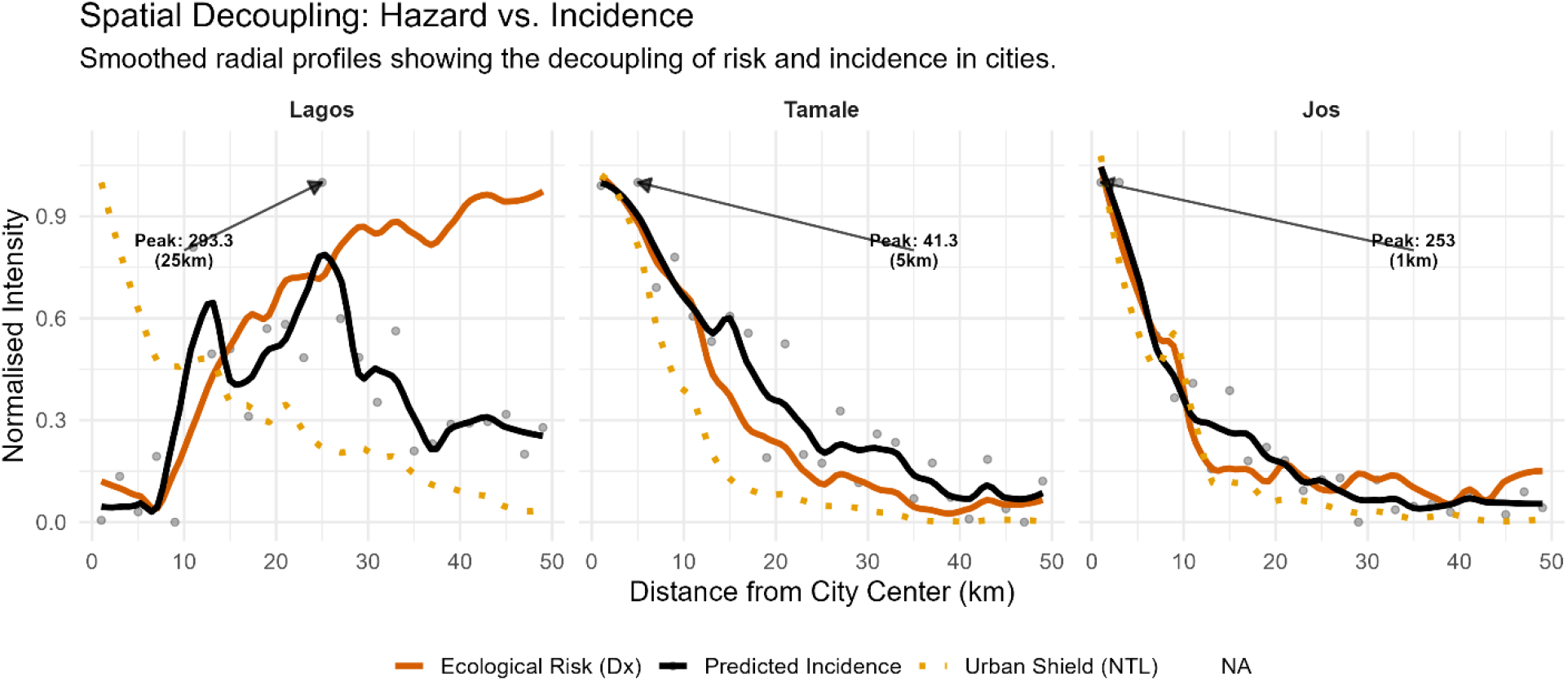
Spatial Decoupling of Hazard and Risk. Radial profiles of Ecological Hazard (Orange), Infrastructure Shield (dotted Yellow), and Predicted Incidence (Black) for representative cities. Lines represent smoothed means (LOESS); arrows indicate the location of peak incidence. **Alt-Text for Figure 8:** A three-panel line graph displaying the radial profiles of ecological hazard, infrastructure shielding, and predicted Lassa fever incidence as distance increases from the centres of Lagos, Tamale, and Jos. The visual starkly contrasts different transmission typologies. In Jos and Tamale, the predicted incidence peaks very close to the urban core (at 1 km and 5 km, respectively) and steadily declines moving outward. In contrast, the Lagos panel demonstrates profound spatial decoupling: predicted incidence remains heavily suppressed by the infrastructure shield near the city centre, only rising after 7 kilometers to reach its peak in the distant peri-urban fringe at approximately 25 kilometers from the core.

Urban typology significantly predicted the spatial location of peak incidence (Kruskal-Wallis *χ*^2^ = 13.34, df = 2, *p* = 0.001 ):

- **Towns and Secondary Cities:** In smaller urban centres (pop < 300,000), the shield is effectively absent. Ecological hazard and human incidence overlap spatially, resulting in intense transmission concentrated in the urban core (n = 61, median = 1 km, IQR = 1–5 km).
- **Regional Cities:** In intermediate cities, a transitional profile is observed. Risk begins to displace from the centre, resulting in a bridge typology between rural towns and major metropolises (n = 23, median = 5 km, IQR = 1–7 km).
- **Large Cities:** In major metropolitan areas (pop > 1 million), the risk profile shifts significantly outward. The median peak incidence is displaced to 8 km from the centre, with a wide variance (n = 20, IQR = 3–14 km) that reflects the heterogeneity between unshielded inland cities (e.g., Ibadan) and strongly shielded developed cities (e.g., Lagos).

### 3.4. The Burden of Infection and Surveillance Gaps

Accounting for this spatial decoupling of hazard and risk yields revised regional burden estimates. Previous models assuming lifelong immunity and rural-dominated transmission estimated approximately 900,000 annual infections across the region. By incorporating the urban shield and correcting for seroreversion (λ= 0.03 year^-1^), I estimate the annual burden of LASV infection to be approximately 2.6 million infections (Sensitivity Range: 0.9–4.4 million, depending on immunity duration) (Figure 9).

**Figure 9.**
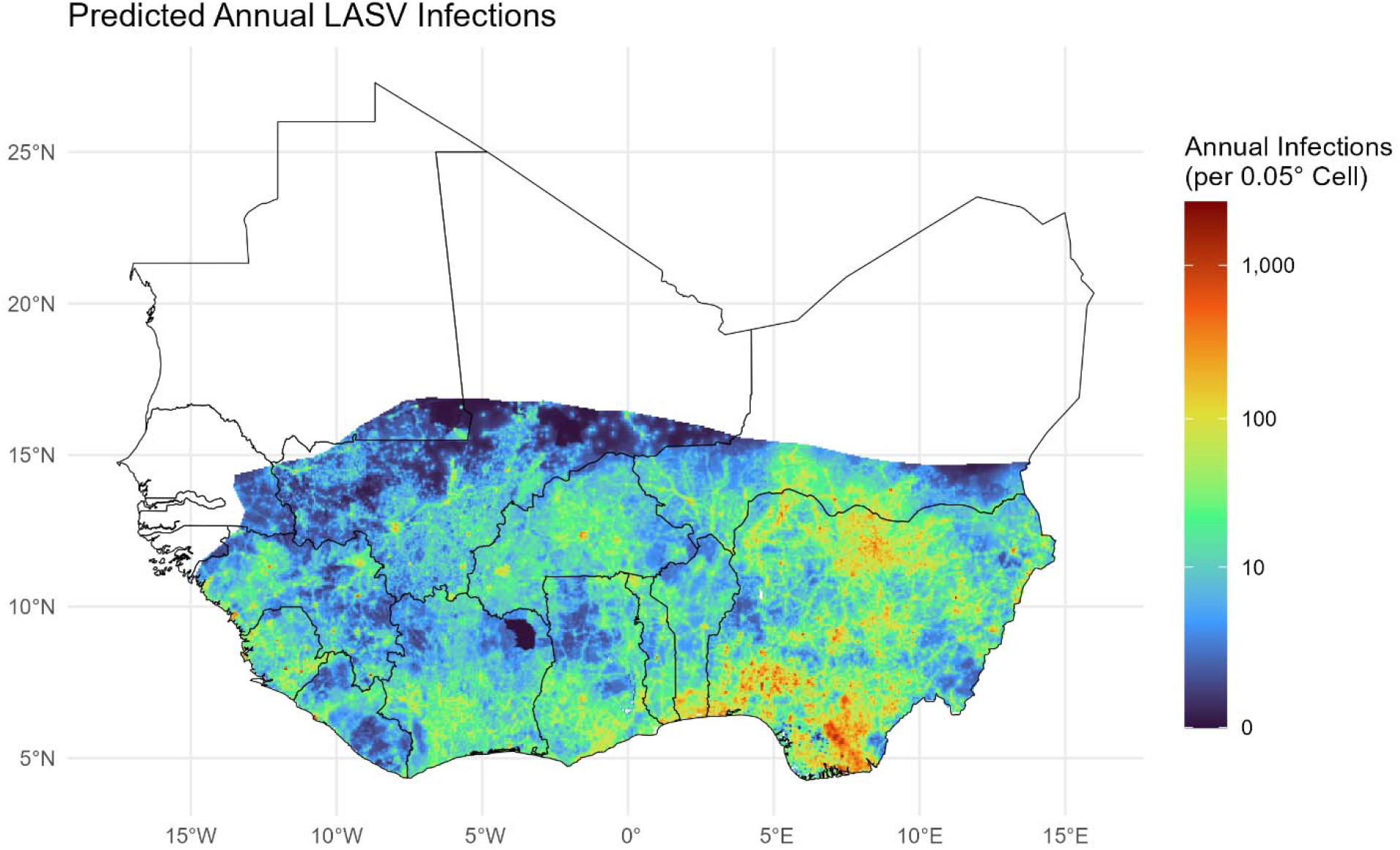
Predicted Annual Incidence of Lassa Virus Infection. Estimates account for seroreversion (λ= 0.03) and the urban shield effect. Scale is log-transformed. **Alt-Text for Figure 9:** A spatial map of West Africa displaying the predicted annual incidence of Lassa virus infections on a log-transformed scale. The visual highlights distinct transmission hotspots clustered in highly populated areas, specifically across Southern Nigeria, the broader coastal region of West Africa, Eastern Sierra Leone, Southern Guinea, and Liberia. Additionally, the map reveals intense pockets of high annual infections in the southern regions of Benin and Togo.

Notably, while Nigeria bears the highest absolute 295 burden due to its population size (1.5 million annual estimated infections), the per-capita infection rates are highest in the Mano River Union region (Guinea: 7.6 per 1000; Sierra Leone: 7.2 per 1000 and Liberia: 6.1 per 1000) and the silent countries of Benin (7.1 per 1000) and Togo (5.6 per 1000) (Table 1).

**Table 1.** Estimated Annual Burden of Lassa Virus Infection by Country. Estimates derived from the Full Custom (Biotic + Shielded) model assuming a seroreversion rate of *λ* = 0.03 year^-1^. Countries are ordered by infection rate (per 1000 people).

| Country | Infections (Thousands) | Rate (per 1000 people)<sup>1</sup> |
| --- | --- | --- |
| Guinea | 100.2 | 7.6 |
| Nigeria | 1,507.8 | 7.4 |
| Sierra Leone | 61.5 | 7.2 |
| Benin | 88.8 | 7.1 |
| Niger | 163.0 | 6.7 |
| Burkina Faso | 134.9 | 6.5 |
| Liberia | 30.1 | 6.1 |
| Ivory Coast | 153.2 | 5.8 |
| Togo | 46.1 | 5.6 |
| Mali | 110.2 | 5.4 |
| Ghana | 142.9 | 4.5 |
| Mauritania | 4.7 | 0.9 |
| Senegal | 3.2 | 0.2 |
| Guinea-Bissau | 0.1 | 0.1 |

<sup>1</sup>Rate calculated using 2020 WorldPop population estimates constrained to the study region.

Finally, comparing predicted annual infections against reported clinical case counts reveals substantial surveillance gaps (Figure 10). Across the entire region, the overall rank correlation between predicted and observed burden is weak (Spearman’s *ρ* = 0.10, *p* = 0.06). However, geographical stratification demonstrates this is heavily driven by regional reporting variance. In the Mano River Union, where historical research is robust, predictions align more closely with historical case counts (AUC = 0.66, *ρ* = 0.24). Conversely, in Nigeria, the correlation degrades (AUC = 0.55, *ρ* = 0.07) as the model identifies numerous silent districts, areas with top-quartile predicted incidence but zero reported cases.

**Figure 10.**
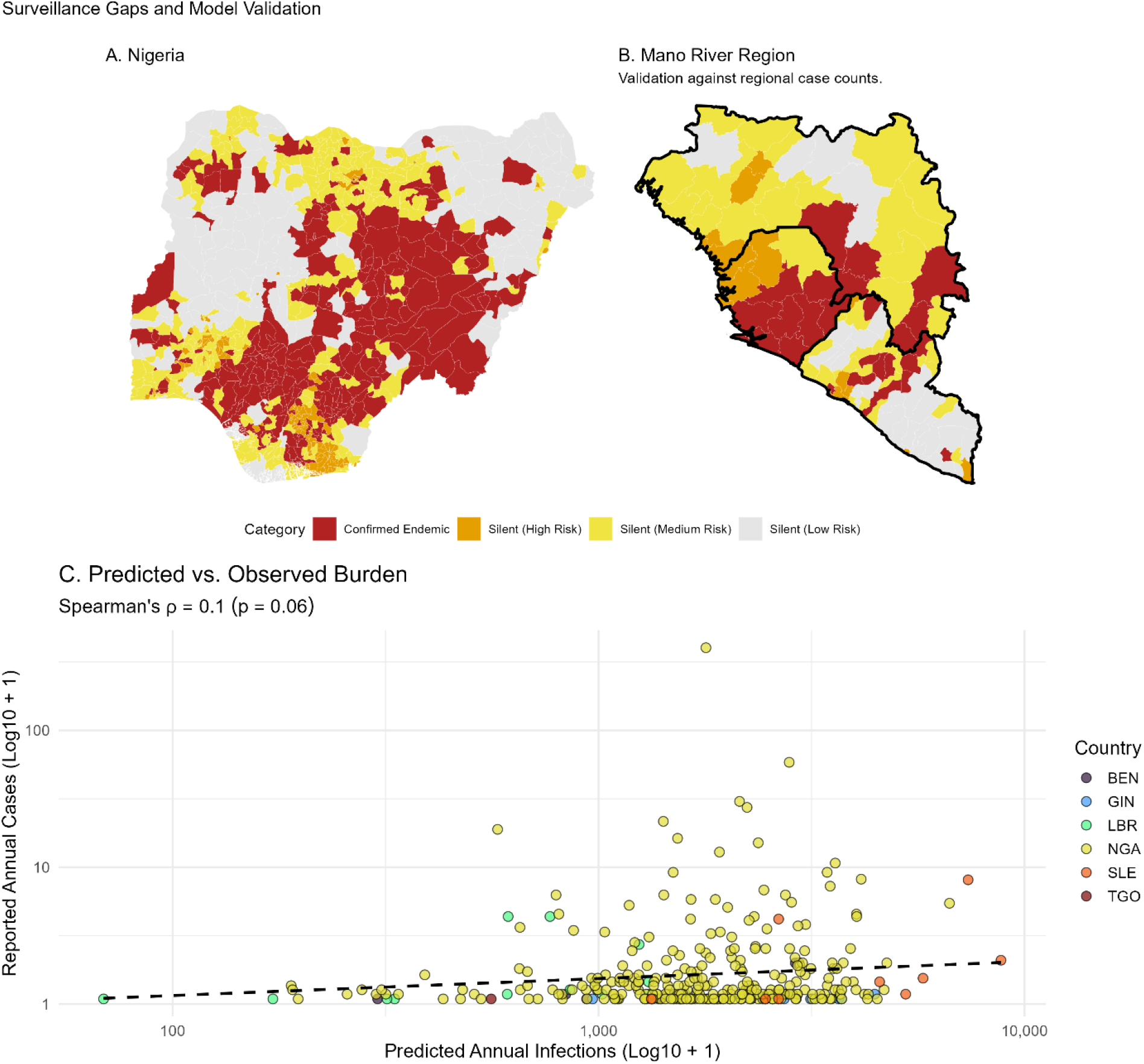
Surveillance Gaps and Model Validation. (A) Nigeria: Risk stratification of LGAs with zero reported cases (NCDC data). (B) Mano River Union: Validation against historical case counts (Moore et al.). Red districts indicate confirmed endemic presence; Orange and Yellow districts represent ‘silent’ areas with high predicted risk but zero reported cases. (C) Scatterplot of predicted annual infections versus reported annual cases across all sub-national districts, coloured by country, illustrating the impact of zero-reporting on overall correlation. **Alt-Text for Figure 10:** A three-panel figure assessing surveillance gaps by comparing model predictions against reported clinical cases. Panel A is a risk map of Nigeria, visually highlighting silent areas, regions with high to medium predicted risk but zero reported cases concentrating heavily in the south and central regions, contrasting with confirmed endemic clusters in the south and the Jos Plateau. Panel B maps the Mano River Union, displaying a much denser network of confirmed endemic districts across Sierra Leone, Guinea, and northern Liberia, though significant high- and medium-risk silent zones remain. Panel C is a scatterplot of predicted infections versus reported cases. The visual is dominated by a dense, horizontal cluster of data points lying flat along the zero-mark of the y-axis (zero reported cases) despite spreading widely across the x-axis (predicted infections), clearly illustrating how massive under-reporting suppresses the overall statistical correlation.

## 4. Discussion

### 4.1. Re-defining the Reservoir Niche: The Urban Blind Spot

Previous spatial models have consistently predicted the exclusion of the *M. natalensis* reservoir from heavily human-modified landscapes [1]. By integrating multi-species interactions into an IMSOM framework, I reveal that the reservoir’s realised niche is significantly constrained by the wider rodent community structure, resulting in high predicted suitability in the peri-urban fringes of major West African cities (Figure 2). This urban blind spot in earlier models likely stems from the substantial rural sampling bias identified in historical trapping data (Figure 1), aligning with recent field surveys documenting *M. natalensis* persisting alongside invasive species in human settlements [15,20,21].

This urban tolerance extends to the pathogen itself. The current model indicates that LASV hazard is sustained in dense peri-urban transition zones where the reservoir and invasive competitors (*R. rattus, M. musculus*) co-occur. While the exact mechanisms of this interaction, whether competitive exclusion or stable co-existence, require finer-scale local study [20,21], the regional epidemiological implication is clear: active surveillance must encompass these expanding peri-urban agricultural belts.

### 4.2. The Socio-economic Shield: A Mechanism for Non-Linear Risk

A limitation of prior spatial models is their reliance on linear, density-dependent transmission assumptions, which project biologically implausible contact rates in megacities [22]. The identification of a Socio-economic Shield (Figure 7) demonstrates that urban infrastructure decouples reservoir presence from human infection. Nighttime Lights (NTL) effectively proxies this transition in housing quality and human behaviour [23]. Consequently, shielded city centres remain relatively LASV-free despite high biological hazard, while unshielded peri-urban fringes suffer intense transmission pressure (Figure 8).

### 4.3. Resolving the Burden and Silent Districts

This biotically-informed model estimates approximately 2.6 million annual LASV infections across West Africa (Table 1). This exceeds prior estimates of 300,000 to 900,000 [1,2], primarily due to incorporating a realistic seroreversion rate (*λ* = 0.03 year^-1^) rather than assuming lifelong immunity. Notably, previous modelling has estimated up to 4.3 million annual infections when allowing for seroreversion [1]. The more conservative 2.6 million burden estimated here reflects the suppressive effect of the Socio-economic Shield, which recalibrates the spatial intensity of transmission out of high-density urban cores.

However, this high infection burden does not imply millions of clinical LF cases. Applying the hospital-derived Case Fatality Rate (∼18%) would suggest >460,000 annual deaths, contradicting regional all-cause mortality data. Instead, this highlights that the vast majority of infections cause asymptomatic or paucisymptomatic seroconversion. The implied Infection Fatality Rate (IFR) is approximately 0.2% to 0.4%, consistent with early historical estimates [2].

This under-reporting is strongly supported by the spatial validation. While predictions align moderately well with clinical data in the highly-surveilled Mano River Union, the model identifies numerous silent districts in Nigeria, Benin, and Togo (Figure 10). These top-quartile incidence areas with zero reported cases represent quantifiable surveillance gaps where the virus likely circulates undetected due to a lack of diagnostic capacity, rather than an absence of biological hazard.

### 4.4. Implications for Control and Elimination

These distinct urban risk typologies require stratified LF control. In towns where the socio-economic shield is weak, vector control and housing improvements must target the urban core. Conversely, in megacities (e.g., Lagos, Abidjan), resources should be prioritised towards the peri-urban fringe (8–20 km radius) where high reservoir density and unshielded housing collide. Furthermore, while *R. rattus* occupancy strongly predicts viral prevalence (Figure 4), it likely signals highly disturbed peridomestic habitats rather than direct LASV transmission. Consequently, indiscriminately targeting all rodents may yield diminishing returns [24]; control must be ecologically targeted to the specific commensal niche of the reservoir.

### 4.5. Limitations

This study is subject to inherent macro-ecological limitations. Predictions at a 0.05^°^ resolution cannot resolve fine-scale household heterogeneity in infrastructure. Furthermore, the Socio-economic Shield represents a semi-mechanistic constraint rather than a purely empirical derivation. Because historical community serosurveys are almost entirely restricted to rural populations, the urban calibration curve is heavily reliant on informative synthetic priors designed to prevent the extrapolation of rural contact rates into concrete-dominated city centres. While grounded in the historical absence of urban outbreaks, this shield remains a theoretical construct requiring rigorous field validation. Additionally, *M. natalensis* was treated as a single taxonomic unit, though mitochondrial lineages vary in LASV competence [25]. Finally, assuming statistical independence between host (**D_M_**) and pathogen (**D_L_**) layers during uncertainty propagation, despite a moderate spatial correlation (*r*= 0.54), means the derived uncertainty bounds should be interpreted as conservative.

Future work must address the critical data gaps exposed by this analysis. Most urgently, systematic human serosurveys must be conducted within the silent districts and across the peri-urban-to-urban gradient to empirically test the Socio-economic Shield hypothesis. Furthermore, accurate quantification of the asymptomatic fraction via community-based cohorts is essential to refine these burden estimates.

## 5. Conclusion

Lassa fever possesses the biological potential to become a peri-urban disease. By moving beyond simple climatic correlations and explicitly modelling the biotic and socio-economic filters of spillover, this study exposes a cryptic burden of infection extending well beyond currently recognised endemic zones. As West Africa urbanises, the protective shield of infrastructure will likely lag behind the expansion of the reservoir’s commensal niche. Bridging this gap through targeted surveillance in silent districts and robust investment in peri-urban housing quality, is a critical public health priority.

## Data Availability

No primary data was generated as part of this study. All data used is available in the associated github repository.

https://github.com/DidDrog11/lassa-bridging-reanalysis

## Statements and Declarations

### Author Contributions

David Simons: Conceptualization, Methodology, Software, Validation, Formal analysis, Investigation, Data Curation, Writing – Original Draft, Writing – Review & Editing, Visualization.

### Data Accessibility Statement

The data and code underlying this article are available in the GitHub repository: https://github.com/DidDrog11/lassa-bridging-reanalysis

### Use of AI Tools

During the preparation of this work, the author used generative AI tools (Google Gemini) to improve the readability, flow, and structural organisation of the manuscript. Additionally, these tools were utilised to assist with error checking, debugging, and commenting of the R analysis scripts. The author takes full responsibility for the content, accuracy, and scientific integrity of the final publication.

### Ethics Statement

This study utilised secondary data sources (published literature, GBIF, and aggregated surveillance reports). As such, no ethical approval was required for primary human or animal subject research.

