## Supplementary Material for "The Socio-economic Shield Limits Lassa Virus Spillover in Urban West Africa"

David Simons

2026-04-14

### 1. Supplementary Methods

#### 1.1. Environmental Predictor Assembly

I reconstructed the environmental predictor stack used in previous forecasting efforts, updating the temporal window to 2001–2025 to reflect contemporary conditions.

- **Climate and Topography:** Mean daily temperature and precipitation were derived from WorldClim 2.1 bioclimatic variables [1]. Elevation (SRTM) was included to account for altitudinal constraints on rodent distribution [2].
- **Seasonality and Extremes:** To account for the strong seasonal forcing of *M. natalensis* demography (which typically peaks in the dry season) and viral shedding, I processed 25 years of monthly CHIRPS precipitation and MODIS NDVI data [3]. Indices of Constancy ($P_{c}$) and Contingency ($P_{m}$) were calculated to explicitly proxy the environmental stability required to sustain high-density reservoir populations year-round, addressing the limitation of static presence-only data.
- **Land Cover Stability:** I analysed 25 years of MODIS Land Cover (MCD12Q1) data to calculate the inter-annual stability of land cover classes [4]. The Urban/Built-up ($LC_{13}$) and Cropland ($LC_{12}$) density layers were prioritised for the final model to explicitly test the hypotheses of competitive exclusion in cities and reservoir preference for agricultural zones.
- **Anthropogenic Factors:** Human population density for 2020 was obtained from the WorldPop constrained dataset and log-transformed to account for extreme skew [5]. Additionally, I incorporated Nighttime Lights (VIIRS 2020 Median Masked) as a proxy for electrification and housing quality, distinguishing between less developed high-density city outskirts (high population, low light) and developed urban cores (high population, high light) [6].

#### 1.2. Integrated Multi-Species Occupancy Model

Host occurrence data were stratified into two observation models to account for heterogeneity in sampling effort. Structured surveys (Source 1) were treated as detection/non-detection histories, with detection probability (p) modelled as a function of sampling effort (log-transformed trap nights). Opportunistic records (Source 2) were treated as presence-only data. To facilitate occupancy modelling, 2,000 random pseudo-absences were generated [7]. To prevent these pseudo-absences from artificially designating under-sampled regions as ecologically unsuitable—a common source of error in correlative models—spatial sampling bias was accounted for by estimating a separate, weakly informative detection intercept for opportunistic data, allowing the model to decouple true ecological absence from lack of observer effort.

The ecological process (true occupancy, $\psi$) for species $i$ at site $j$ was modelled as:

$$\text{logit}\left( \psi_{i,j} \right)=\alpha_{i}+\mathbf{X}_{j}\boldsymbol{\beta}_{i}+\eta_{i,j}$$

Where $\mathbf{X}_{j}$ represents the vector of environmental covariates (e.g., climate, land cover, and urbanisation). $\boldsymbol{\beta}_{i}$ represents the species-specific responses to these environmental gradients. The term $\eta_{i,j}$ captures the residual species correlations via a latent factor analysis ($q=2$ factors), enabling the quantification of interspecific covariance ($\boldsymbol{\rho}_{ij}$) independent of shared environmental preferences.

The model was fit using Markov Chain Monte Carlo (MCMC) with 3 chains of 50,000 iterations (burn-in = 20,000, thinning = 10). Convergence was assessed using the Gelman-Rubin statistic (($\hat{R}<1.1$)) and visual inspection of Effective Sample Size (ESS).

While sporadic natural infections of LASV have been reported in invasive species, available evidence suggests they are unlikely to maintain transmission. Analysis of the ArHa dataset indicates that *M. natalensis* exhibits substantially higher active infection rates (PCR positivity: 14.4%, n=4,339) and historical exposure (Seroprevalence: 17.9%, n=2,989) compared to sympatric invasives. *M. musculus* showed low acute infection (PCR: 2.5%, n=80) and negligible seroprevalence 0.16%, n=619). Similarly, *R. rattus* showed low active infection (PCR: 0.6%, n=301) and low seroprevalence (1.0%, n=808). Because these limited detections typically occur in high-transmission environments shared with *M. natalensis*, the IMSOM treats their occurrence as an ecological modifier of the primary reservoir niche rather than an independent hazard.

#### 1.3. SIRS Differential Equations and Steady-State Solution

To estimate human incidence from spatial seroprevalence, I defined a demographic Susceptible-Infectious-Recovered-Susceptible (SIRS) compartmental model. Let $S$, $I$, and $R$ represent the proportion of the human population in the susceptible, infectious, and recovered (seropositive) states, respectively. Assuming a constant population size ($S+I+R=1$), the transmission dynamics are described by the following system of ordinary differential equations:

$$\begin{matrix} \frac{dS}{dt} & =\mu-\lambda_{\text{FOI}}S-\mu S+\lambda R \\ \frac{dI}{dt} & =\lambda_{\text{FOI}}S-\left( \gamma+\mu_{d} \right)I \\ \frac{dR}{dt} & =\gamma I-\left( \lambda+\mu\right)R \end{matrix}$$

where $\mu$ represents the background human demographic turnover (birth and natural death rates), $\lambda_{\text{FOI}}$ is the force of infection, $\gamma$ is the recovery rate from infectiousness, $\lambda$ is the rate of antibody waning (seroreversion), and $\mu_{d}$ represents the disease-specific mortality rate.

The disease-specific mortality rate can be expressed in terms of the infection fatality rate ($f$) as $\mu_{d}=\mu+\frac{f\gamma}{1-f}$. Therefore, the total exit rate from the infectious compartment simplifies to $\frac{\mu+\gamma}{1-f}$.

Because the duration of acute LASV infection is brief (measured in weeks) relative to the human lifespan, the proportion of the population currently infectious at any given time is negligible ($I\approx0$). Consequently, the population can be approximated as $S\approx1-R$.

At endemic steady-state, we set $\frac{dR}{dt}=0$, yielding:

$$\begin{matrix} \gamma I=\left( \lambda+\mu\right)R \end{matrix}$$

Substituting $I$ back into the steady-state equation for the infectious compartment ($\frac{dI}{dt}=0$):

$$\begin{matrix} \lambda_{\text{FOI}}S=\frac{\mu+\gamma}{1-f}I \end{matrix}$$

By substituting $\gamma I$ with $\left( \lambda+\mu\right)R$ and replacing $S$ with $\left( 1-R \right)$, I solve for the steady-state incidence of new infections (the flow from $S$ to $I$):

$$\begin{matrix} \text{Incidence Proportion}=\lambda_{\text{FOI}}\left( 1-R \right)=R\left[ \frac{\left( \mu+\gamma\right)\left( \mu+\lambda\right)}{\gamma\left( 1-f \right)} \right] \end{matrix}$$

By substituting the model-predicted spatial seroprevalence ($\Omega^{*}$) for $R$ and multiplying by the total population count ($N$), we arrive at the final analytical equation utilised in the main text to calculate absolute annual infections per pixel.

#### 1.4. Urban Gradient Analysis and Stratification

To quantify the divergence between biological hazard and realised incidence in urban environments, a spatial gradient analysis was performed across 104 West African settlements. The sample included 12 focal cities representing distinct historical transmission typologies (e.g., Lagos, Tamale, Kenema) and a stratified random sample of 92 additional settlements to ensure regional representativeness. The stratified sampling selected 5 cities per population class per country.

Urban centres were classified based on their estimated 2020 population within a 10 km radius, derived from the constrained WorldPop raster used in the primary incidence modelling. This ensured that classifications reflected the demographic reality of the study period, correcting for the rapid, ongoing urbanisation of West Africa. Classes were defined as: Towns (< 300,000 population), Regional Cities (300,001–1,000,000), and Large Cities (> 1,000,000).

For each settlement, mean values for Ecological Hazard ($\mathbf{D}_{\mathbf{X}}$), the Socio-economic Shield (NTL), and Realised Incidence were extracted in 2 km concentric rings along standardised 50 km radial transects extending from the city centre. To determine whether the spatial location of peak incidence (distance in km from the urban core) differed significantly between urban demographic typologies, a Kruskal-Wallis rank sum test was applied to the extracted radial profiles.

### 2. Supplementary Figures

#### 2.1 Supplementary Figure 1

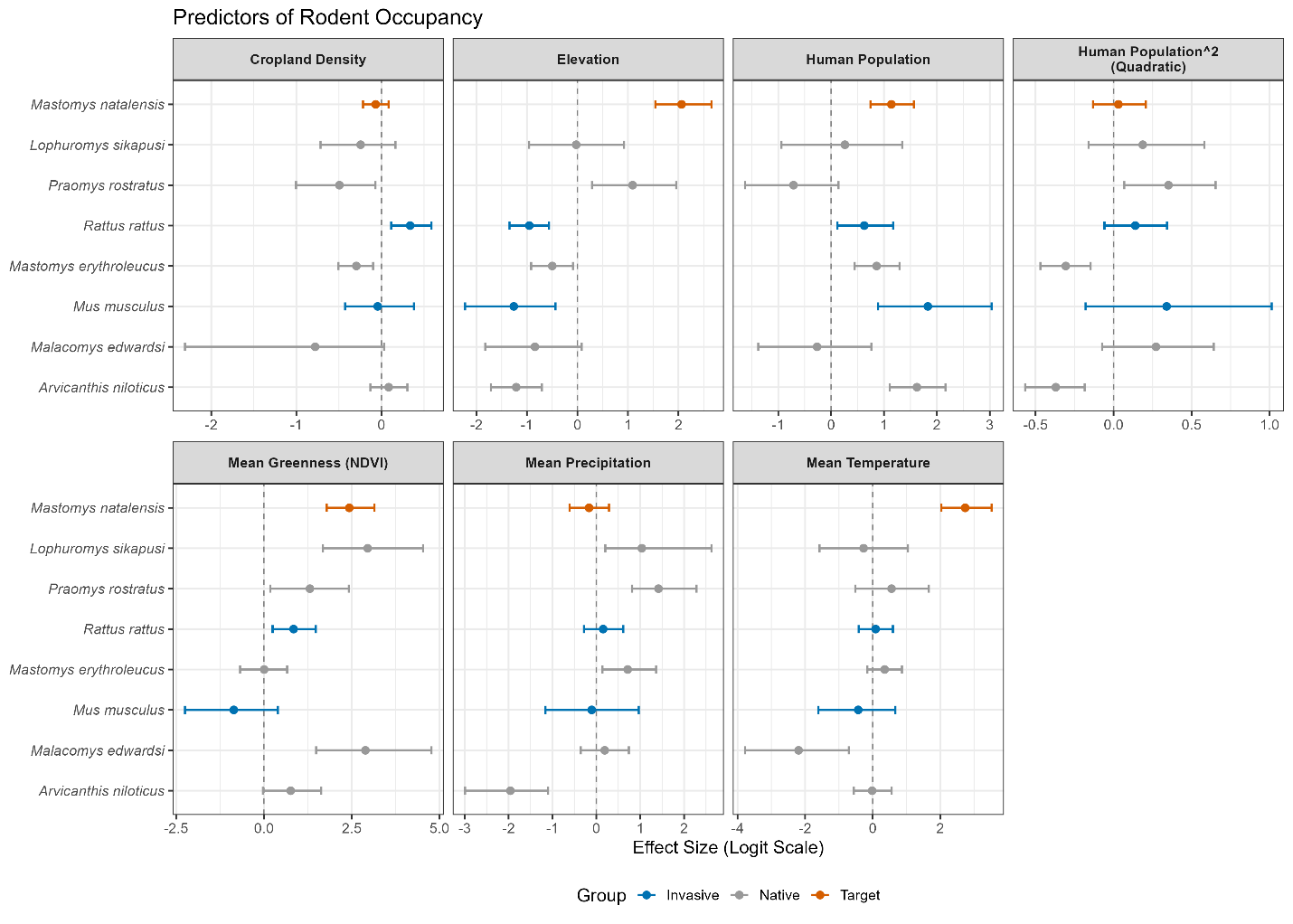

**Supplementary Figure 1.** Species-specific environmental response coefficients from the Integrated Multi-Species Occupancy Model (IMSOM). Posterior mean estimates (points) and 95% credible intervals (lines) for the standardized beta coefficients of eight target rodent species. Predictors include Mean Temperature (Tmu), Mean Precipitation (Pmu), Mean Greenness (Nmu), Elevation (Elev), Cropland Density (LC12), and Human Population Density (Pop and Pop^2). Coefficients are on the logit scale. The primary reservoir, *Mastomys natalensis* (orange), exhibits a distinct positive association with anthropogenic variables (Population Density) compared to forest-specialist species like *Malacomys edwardsi* (grey), which show negative or neutral responses. Invasive competitors (*Rattus rattus*, *Mus musculus*) are shown in blue. Note the strong positive association of *M. natalensis* with Elevation and Mean Greenness, reflecting its broad ecological tolerance across the derived savannah zone.

#### 2.2 Supplementary Figures 2 & 3

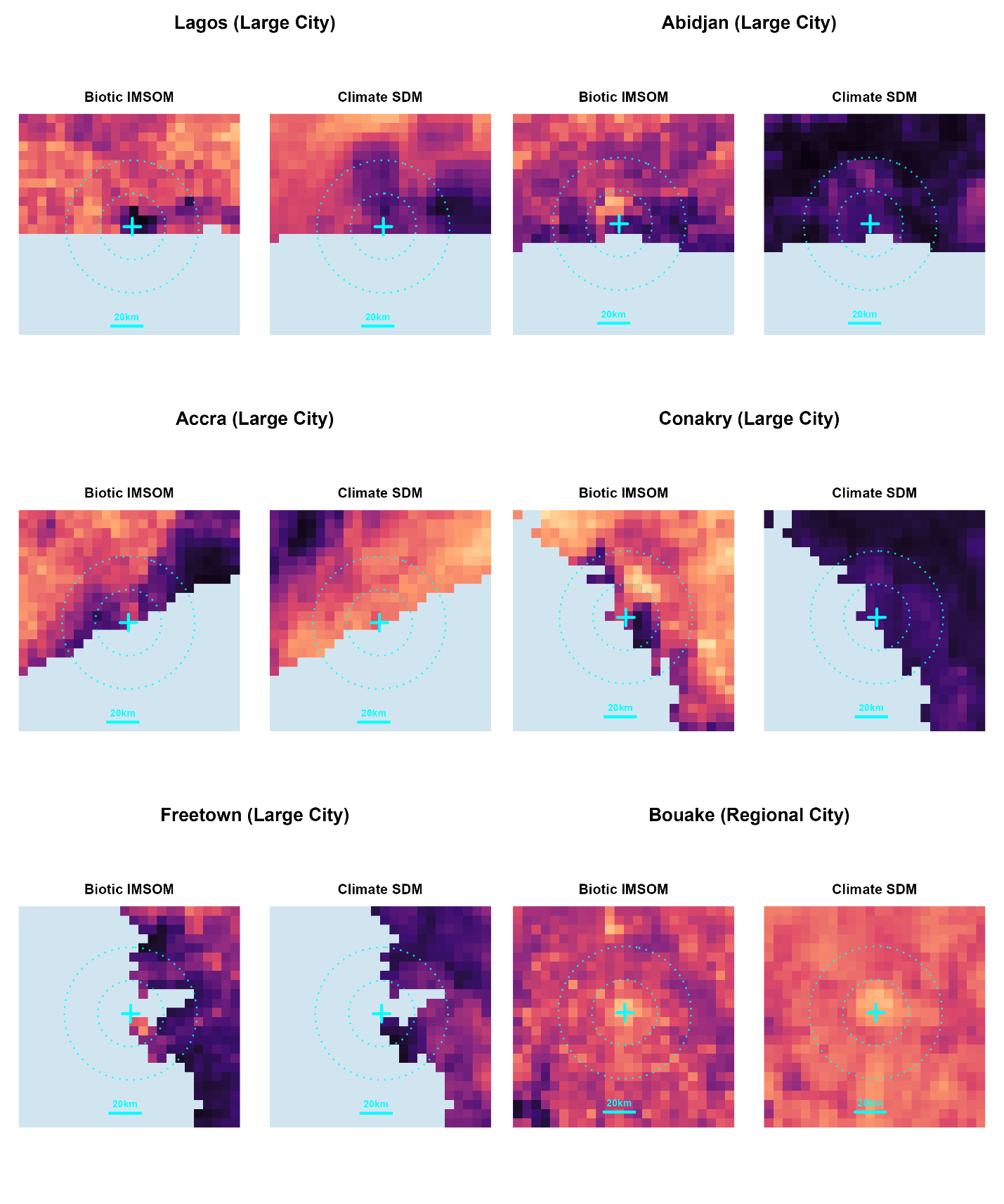

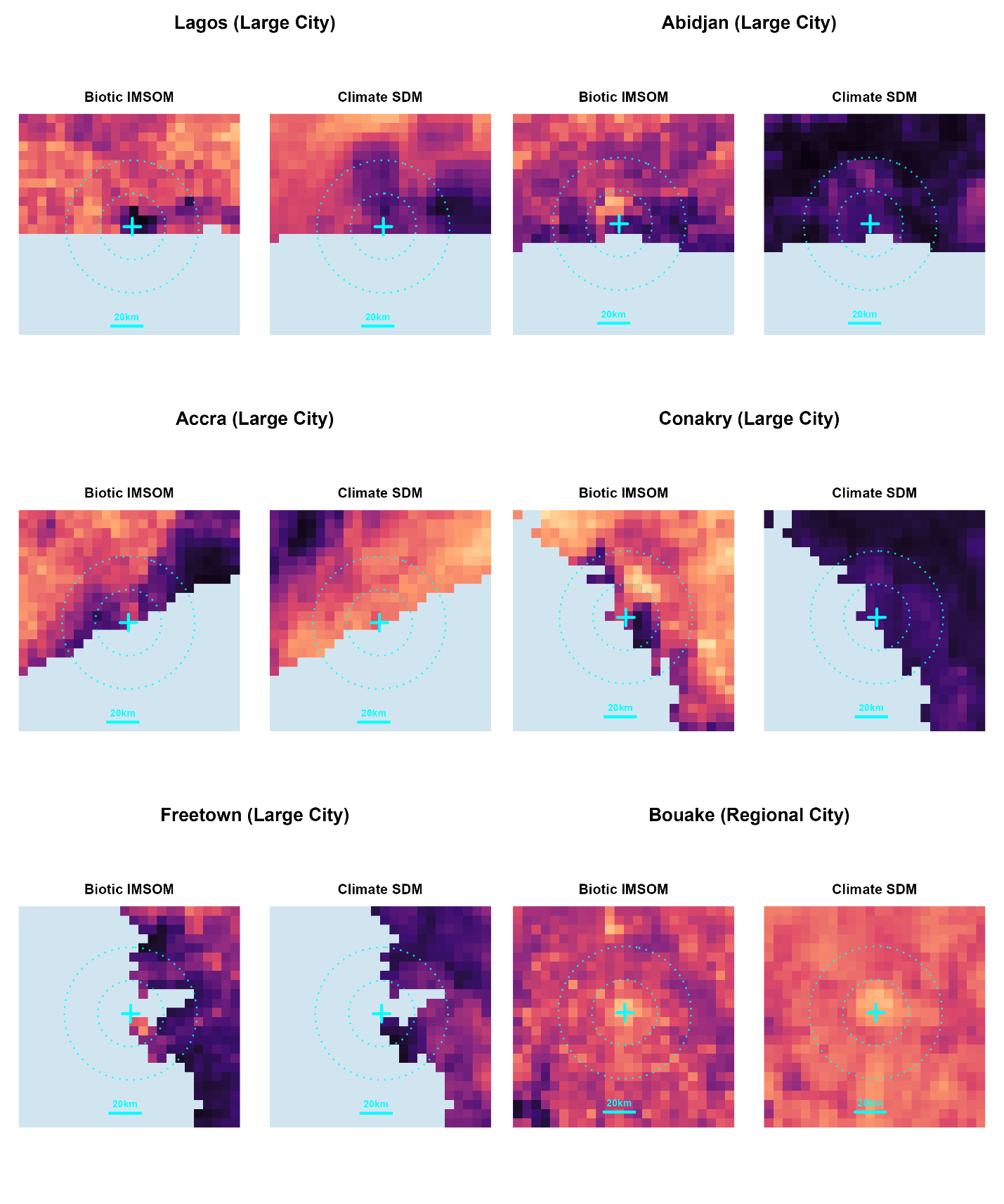

**Supplementary Figures 2 & 3.** Comparative reservoir niche predictions across urban gradients. Spatially explicit occupancy probabilities for *Mastomys natalensis* in and around 12 representative West African settlements, stratified by urban typology. For each location, the panels compare the Climatic SDM (Basinski et al. 2021; right) against the Biotic IMSOM (this study; left). Warmer colours (Magma palette) indicate higher occupancy probability (ψ). Cyan dotted contours indicate radial distances of 20 km and 40 km from the city centre (marked by a cyan cross). Note the consistent divergence in the urban core: the Climatic SDM typically predicts reservoir exclusion (dark voids) in high-density areas, whereas the Biotic IMSOM captures the realised niche, predicting high suitability in the peri-urban fringe (e.g., Lagos, Tamale) and persistence within endemic towns (e.g., Kenema, Ekpoma).

#### 2.3 Supplementary Figure 4

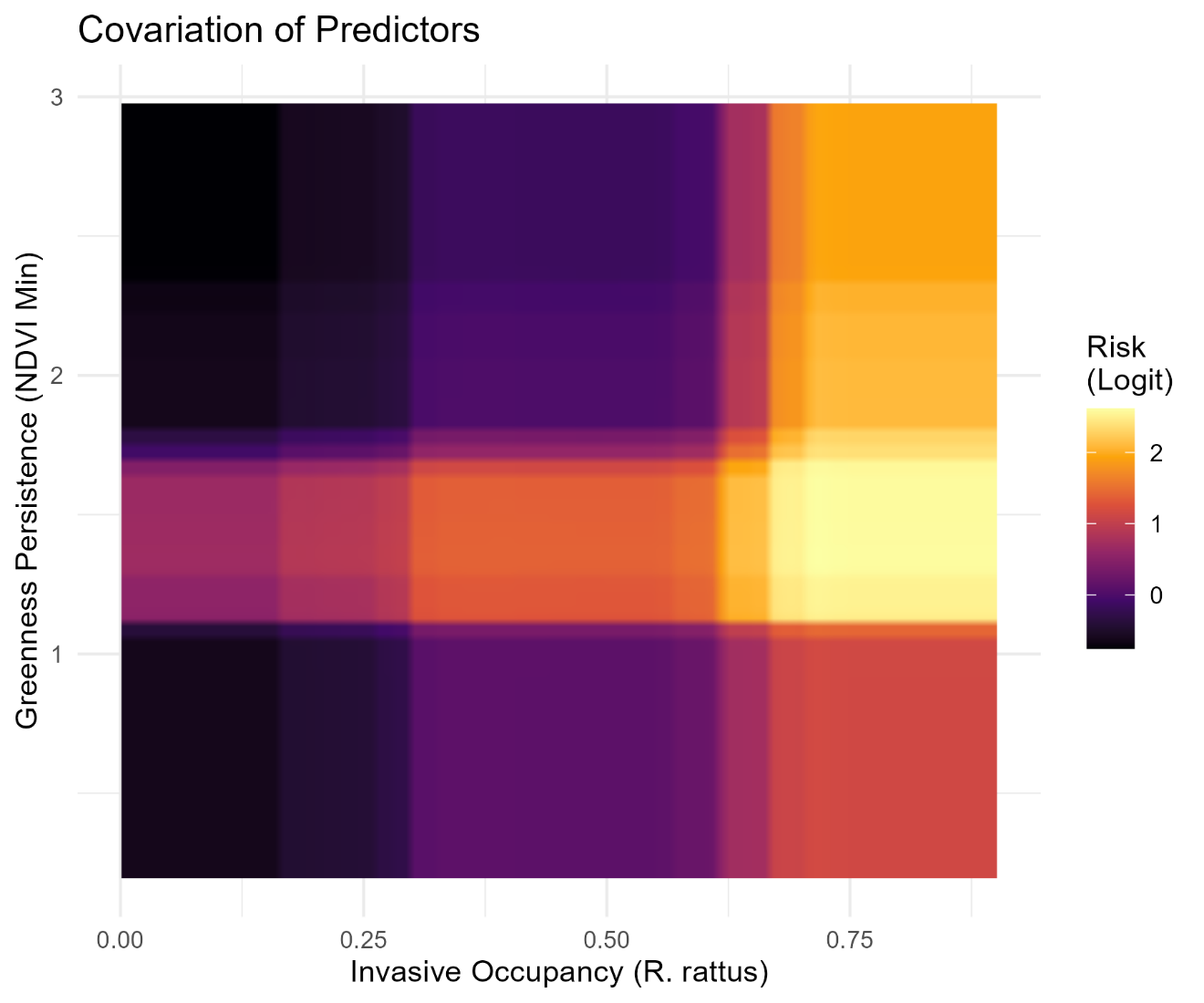

**Supplementary Figure 4.** Bivariate interaction surface of the top environmental and biotic predictors. The joint partial dependence of Lassa virus prevalence on Greenness Persistence (NDVI Min, y-axis) and Rattus rattus occupancy (x-axis). Warmer colours (Inferno palette) indicate higher marginal probabilities of viral presence. The risk surface highlights a specific high-risk typology (brightest yellow) defined by high invasive species occupancy within intermediate vegetation regimes, effectively delimiting the derived savannah zone from the lower-risk deep forest (high NDVI) and arid Sahel (low NDVI).

#### 2.4 Supplementary Figure 5 & 6

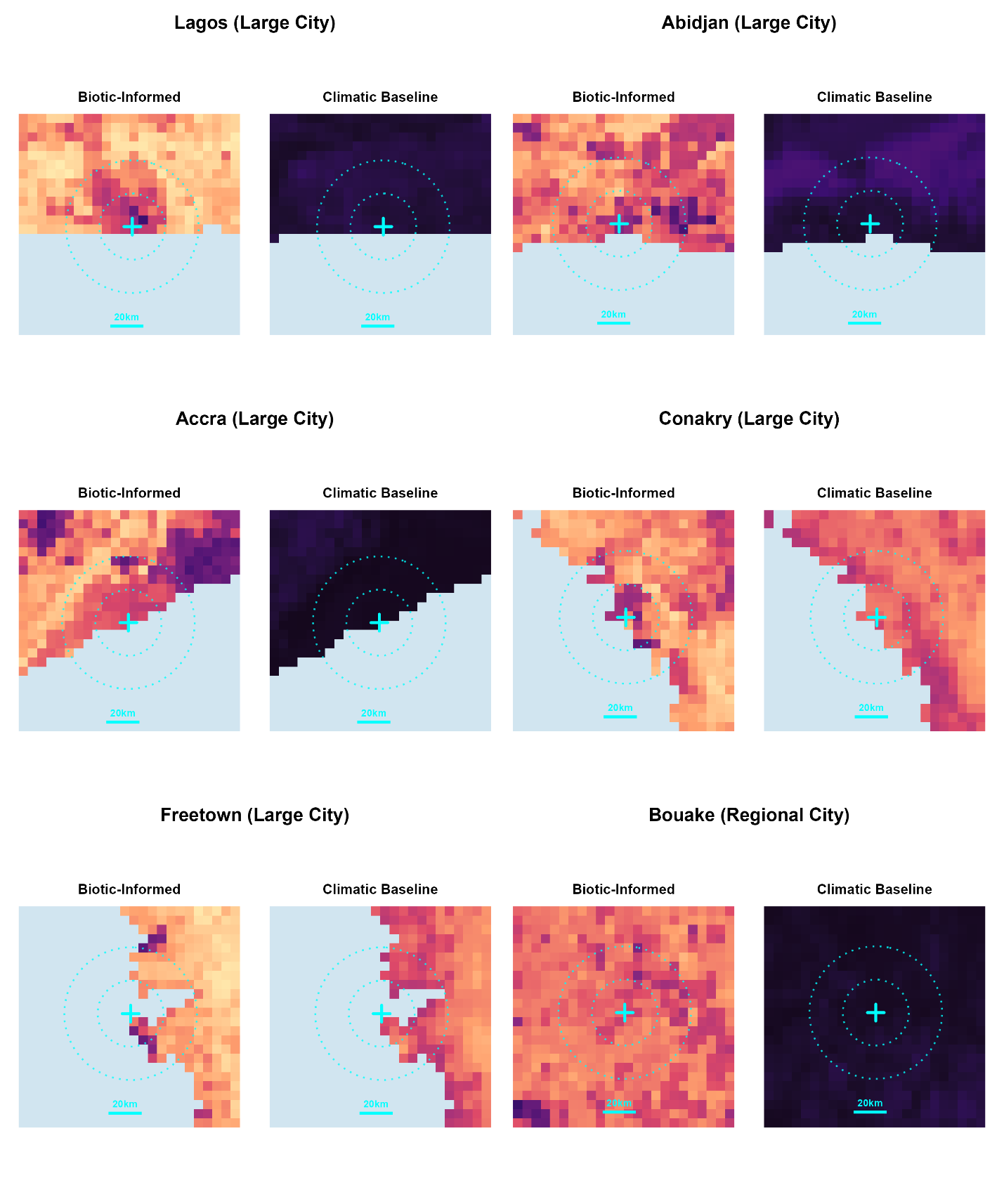

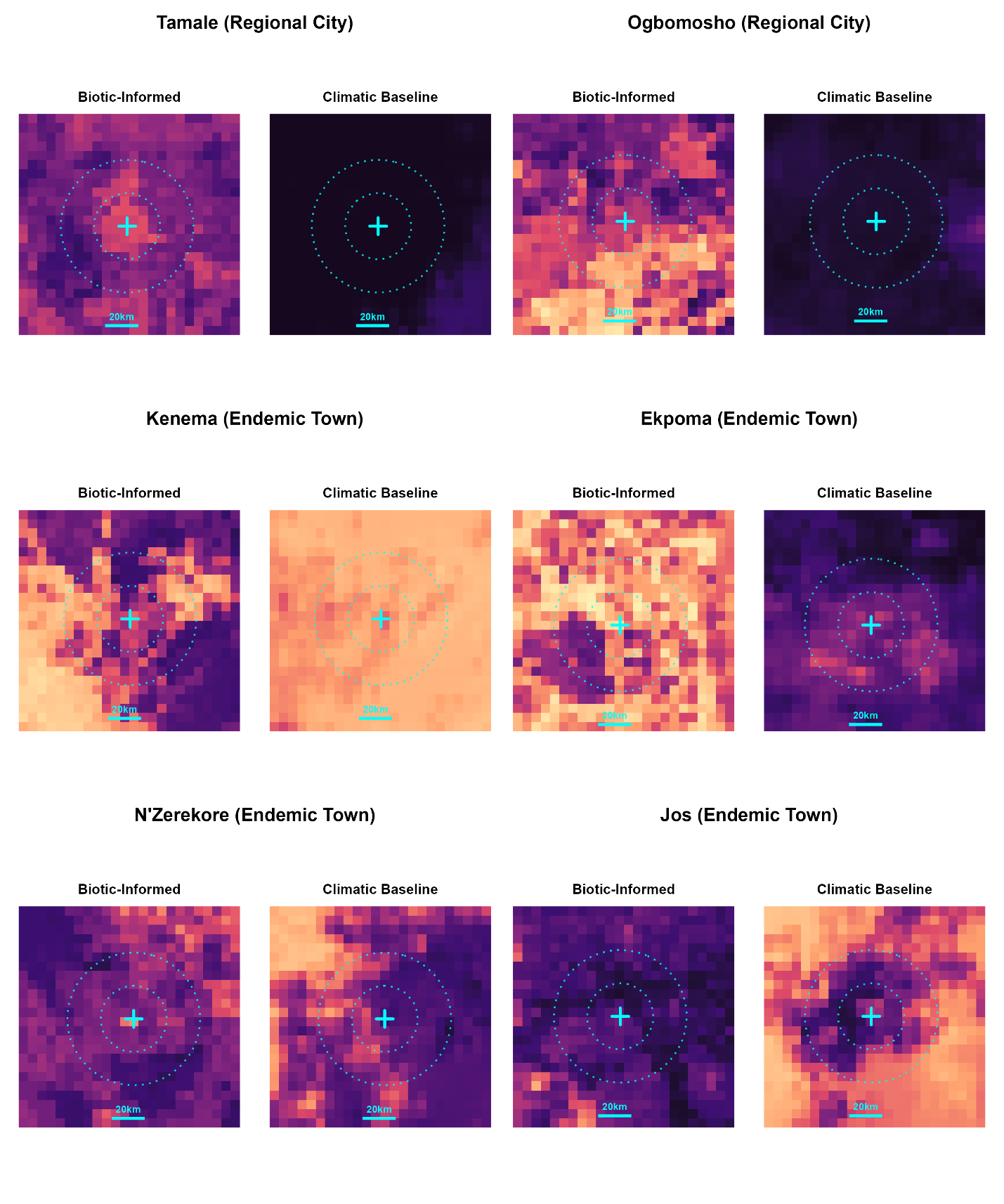

**Supplementary Figures 5 & 6.** Comparative pathogen prevalence predictions across urban gradients. Spatially explicit predictions of Lassa virus prevalence ($\mathbf{D}_{\mathbf{L}}$) in and around 12 representative West African settlements, stratified by urban typology. For each location, the panels compare the Climatic Baseline (Basinski et al. 2021; right) against the Biotic-Informed Model (this study; left). Warmer colours (Rocket palette) indicate higher predicted viral prevalence within the reservoir population. Cyan dotted contours indicate radial distances of 20 km and 40 km from the city centre (marked by a cyan cross). Note that while the Climatic Baseline predicts a sharp decline in risk as human density increases, the Biotic-Informed Model predicts a resurgence of viral risk in the peri-urban transition zones (e.g., surrounding Lagos) where invasive species co-occur with the reservoir, before declining in the shielded urban core.

#### 2.5 Supplementary Figure 7

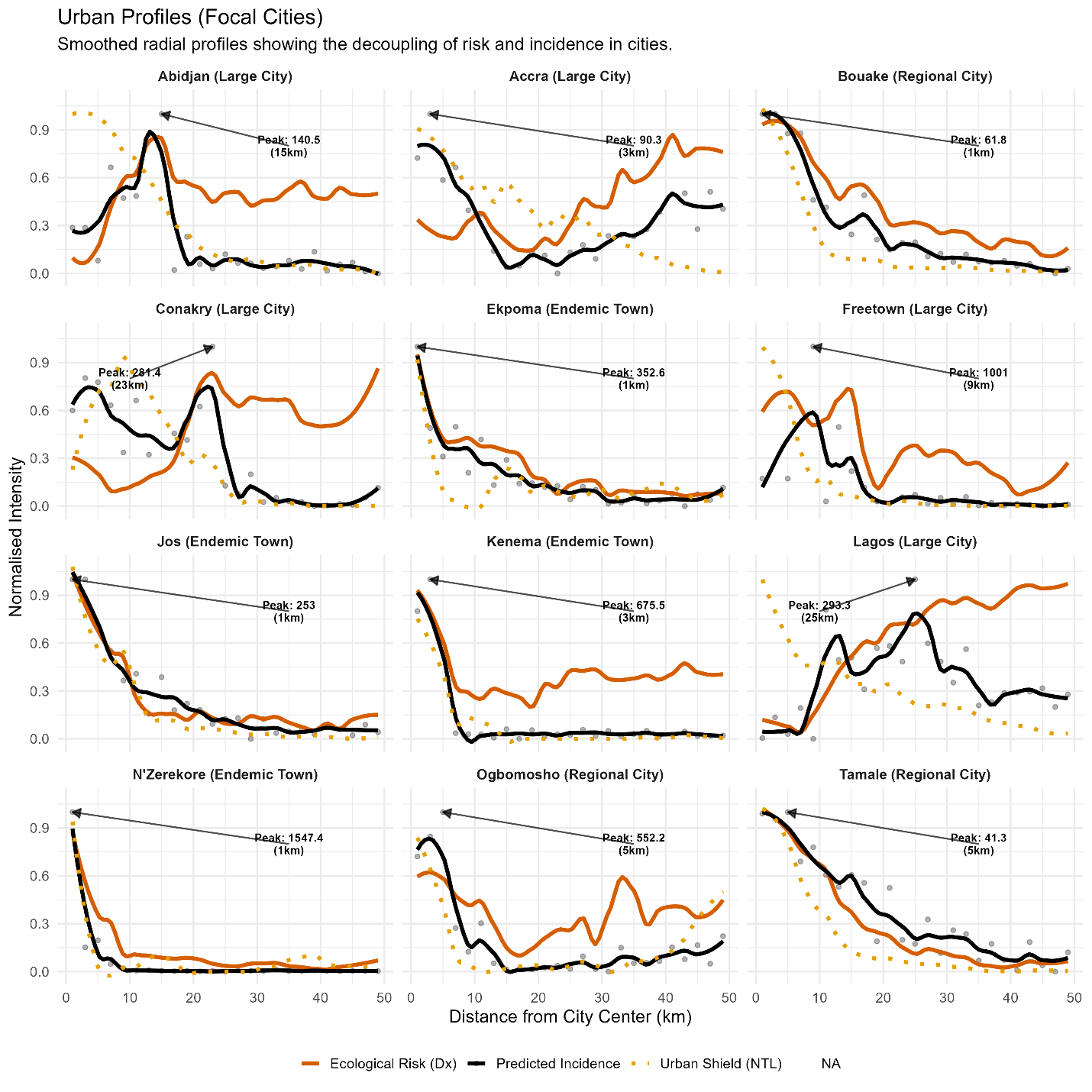

**Supplementary Figure 7.** Radial profiles of spillover risk components across urban typologies. Smoothed radial gradients of Ecological Hazard, Socio-economic Shielding, and Predicted Incidence for 12 focal cities. Normalised intensity (y-axis) is plotted against distance from the city centre (x-axis). The Ecological Hazard ($\mathbf{D}_{\mathbf{X}}$ orange solid line) represents the potential risk driven by reservoir and viral suitability. The Socio-economic Shield (NTL, orange dotted line) proxies infrastructure quality and barrier effects. Predicted Incidence (black line) represents the realised force of infection after shielding is applied. Arrows indicate the spatial location of peak incidence. Note the distinct spatial decoupling in Large Cities (e.g., Lagos, Abidjan and Conakry), where the shield effectively displaces transmission to the peri-urban fringe (>8 km), contrasting with Endemic Towns (e.g., Jos, N’Zerekore, Kenema and Ekpoma) where the shield is absent and incidence spatially overlaps with hazard in the urban core.

### 3. Supplementary Tables

#### 3.1. Supplementary Table 1. Summary of biological and epidemiological data sources.

Data were stratified into four categories: Host Occurrence (used to train the IMSOM), Rodent Testing (used to train the Pathogen BRT), Human Seroprevalence (used for calibration), and Case Reports (used for validation). *N Sites* refers to unique spatial locations. *Context* classifications are based on Dijkstra urbanisation classes.

**Supplementary Table 1: Summary of biological and epidemiological data sources.** Data were stratified into four categories: Host Occurrence (used to train the IMSOM), Rodent Testing (used to train the Pathogen BRT), Human Seroprevalence (used for calibration), and Case Reports (used for validation). N Sites refers to unique spatial locations. Context classifications are based on Dijkstra urbanisation classes.

|  | Study Effort | | | Spatial Context (No. Sites) | | | Species Detections | | |
| --- | --- | --- | --- | --- | --- | --- | --- | --- | --- |
| Data Source | Period | Unique Sites | Positive / Total | City | Town | Rural | M. natalensis*^1^* | Invasive*^2^* | Native |
| **Host Occurrence (IMSOM Training)** | | | | | | | | | |
| Systematic Surveys (ArHa/WA) | 1964-2019 | 677 | — | 158 | 70 | 457 | 630 | 798 | 1,044 |
| Targeted Urban Search | 1980-2021 | 148 | — | 123 | 11 | 13 | 144 | 282 | 77 |
| Opportunistic / GBIF | 1977-2025 | 4,083 | — | 71 | 116 | 3,894 | 844 | 4,415 | 4,301 |
| **Rodent Pathogen Testing (BRT Training)** | | | | | | | | | |
| PCR | 1972-2022 | 54 | 713 | 2 | 12 | 41 | 4,234 | — | — |
| Serology | 1972-2022 | 47 | 459 | 2 | 7 | 38 | 3,949 | — | — |
| Virus Culture | 1972-2022 | 2 | 5 | 0 | 1 | 1 | 51 | — | — |
| **Human Calibration** | | | | | | | | | |
| Human Seroprevalence | 1970-2024 | 197 | 5,262 | 7 | 25 | 165 | — | — | — |
| **Validation Sets** | | | | | | | | | |
| NCDC Surveillance (Nigeria) | 2018-2025 | *^3^*775 | 263 | — | — | — | — | — | — |
| Regional Surveillance (Moore et al.) | 2012-2022 | *^3^*338 | 340 | — | — | — | — | — | — |
| *^1^*For Host Occurrence, values represent site detections (presence). For Rodent Testing, values represent individual animals tested. | | | | | | | | | |
| *^2^*Invasive species: Rattus rattus, Mus musculus. Native species: Malacomys edwardsi, Praomys rostratus, Lophuromys sikapusi, Arvicanthis niloticus, M. erythroleucus. | | | | | | | | | |
| *^3^*For Validation Sets, 'Unique Sites' refers to unique Administrative Level 2 units (e.g., LGAs, Districts) rather than point coordinates. | | | | | | | | | |
